# Intentional and unintentional non-adherence to social distancing measures during COVID-19: A mixed-methods analysis

**DOI:** 10.1101/2021.05.04.21256444

**Authors:** Yolanda Eraso, Stephen Hills

**Author notes:** Corresponding author (YE). **Authors contribution: Conceptualization:** Yolanda Eraso, Stephen Hills, **Formal analysis:** Yolanda Eraso, Stephen Hills, **Methodology:** Yolanda Eraso, Stephen Hills, **Project administration:** Yolanda Eraso, Stephen Hills, **Writing – original draft:** Yolanda Eraso, Stephen Hills, **Writing – review & editing:** Yolanda Eraso, Stephen Hills.

## Abstract

Social distancing measures implemented by governments worldwide during the COVID-19 pandemic have proven an effective intervention to control the transmission of SARS-CoV-2. There is a growing literature on predictors of adherence behaviours to social distancing measures, however, there are no comprehensive insights into the nature and types of non-adherence behaviours. To address this gap in the literature, we studied non-adherence in terms of counts of infringements and people’s accounts on their behaviours in a representative sample of North London residents. We focused on the following social distancing rules: keeping 2 mts. distancing, meeting family and friends, and going out for non-essential reasons.

A mixed-methods explanatory sequential design was used comprising an online survey (1^st^ – 31^st^ May 2020) followed by semi-structured in-depth interviews held with a purposive sample of survey respondents (5^th^ August – 21^st^ September 2020). A negative binomial regression model (quantitative) and Framework Analysis (qualitative) were undertaken.

681 individuals completed the survey, and 30 individuals were interviewed. We integrated survey and interview findings following three levels of the Social Ecological model: individual, interpersonal and community levels. We identified non-adherence behaviours as unintentional (barriers beyond individual’s control) and intentional (deliberate decision). Unintentional adherence was associated with and reported as emotional inability to stay at home, lack of controllability in keeping 2 mts. distancing, social responsibility towards the community and feeling low risk. Intentional non-adherence included individual risk assessment and decision-making on the extent to following the rules, support from friends, and perceived lack of adherence in the local area. Our findings indicate that unintentional and intentional non-adherence should be improved by Government partnerships with local communities to build trust in social distancing measures; tailored messaging to young adults emphasising the need of protecting others whilst clarifying the risk of transmission; and ensuring COVID-secured environments by working with environmental health officers.

## Introduction

On March 23, 2020, the UK government announced a new set of mitigation measures to slow the spread of COVID-19. New regulations required the public to observe social distancing (SD) by staying at home and only leaving to exercise once a day, to shop for essential items, to seek and provide medical assistance, and to travel to work if this was not possible from home. People were asked to minimise the time spent outdoors and to keep a minimum distance of 2 mts. away from others outside their household. In addition, a shielding policy for extremely vulnerable people as well as self-isolation measures for those with symptoms were also introduced [1].

There is a burgeoning literature and data available on the effectiveness of government restrictive measures and on people’s adherence to the rules. Evidence indicates that restriction of movement and the closure of schools and business have proved effective in reducing the transmission of the virus in several countries including the UK [2]. Government data on cases, for example, show that soon after the introduction of the second ‘lockdown’ in the UK (5^th^ November 2020) the number of people who tested positive for COVID-19 dropped from a peak of 33,470 cases on 12^th^ November to 11,299 cases on 24^th^ November 2020 [3]. However, after restrictions were eased four weeks later on 2^nd^ December 2020, cases rose to a new daily peak of 68,053 on 8^th^ January 2021, requiring the UK to enter a third national lockdown on 6^th^ January 2021 [3].

According to recent surveys, people’s adherence to SD measures in the UK reflects variations informed by different timeframes and measures considered: for example, data have shown higher levels of compliance in the first months of the full ‘lockdown’ and a decline towards the end of that period (May) and during the ‘relaxation’ of measures in the summer [4–6]. These levels were measured according to the extent participants followed Government rules and scores for ‘completely/nearly all the time’, which have the limitation of relying on people’s understanding of the rules. In addition, adherence to SD rules has measured different types of protective behaviours (avoiding crowds and social events, keeping 2 mts. distance, meeting friends and family, not self-isolating with symptoms, and shopping for non-essentials) [6–8].

Arguably, the nature of these behaviours are different in more than one sense: some were voluntary (self-isolation/quarantine, ‘staying at home’); some were a personal choice (hygiene); some required new habits (keeping 2 mts. distancing); some were enforced by law (the police having the right to act and issue fines in large gatherings in public places, house party or a crowded shop if physical distance was not observed); some lasted for different periods (‘staying at home’ was discontinued whilst hygiene recommendations and keeping 2 mts. distance continued). In this sense, UK surveys have identified different demographic and psycho-social explanatory factors (such as gender, age, socio-economic status, knowledge, and vulnerability amongst others) for a range of behaviours to which we will return in our discussion.

Evidence suggests that one specific measure to which people were less adherent was self-isolation if the person had symptoms of COVID-19, which according to the CORSAIR study [9], adherence has been significantly low (18%). Differences in the ability of people to comply with these rules depends on the degree of controllability that individuals have on their behaviour. Thus, for many, without Government financial support to self-isolate, the prospect of losing income or risking employment is likely to discourage compliance. Therefore, individual-psychological factors may not be enough to explain decisions to not adhere to SD rules, and the influences of interpersonal, community and environmental conditions, which are constructs germane to the Social Ecological Model [10] can provide a useful framework for achieving a better understanding on non-adherence. Related to this, there is a need to distinguish between whether non-adherence behaviours during mitigation measures and relaxation were intentional (i.e. the result of conscious, deliberate decision-making due to attitudes, beliefs or priorities) or unintentional (unable to comply due to lack of personal ability, environmental controllability or lack of understanding about the guidelines) [11]. Understanding these different types of non-adherence, their associated predictors alongside people’s perceptions, would be useful in tailoring interventions to improve adherence.

Our previous quantitative research [11] modelled factors predictive of whether or not an individual failed to adhere to SD rules over a two-week period, and from the perspective of non-adherence to all SD rules and intentional non-adherence. Results indicated that explanatory factors differed, whereby non-adherence to all SD rules was more associated with vulnerability and control over SD, whereas intentional non-adherence was more associated with intention and anti-social psychological factors. These factors allowed us to provide recommendations to tackle the distinct behaviours of unintentional and intentional non-adherence. The use of a binary measure of non-adherence to all SD rules, however, was limited in variability whereby 92.8% of participants did not adhere to all SD rules. The present study further extended the analysis of non-adherence to SD rules by using a mixed-method design whereby the quantitative phase measured and modelled counts of infringements (to all rules and intentional rule-breaking) to better reflect the complexity and variability of non-adherence behaviours. Further to using a more nuanced outcome variable, the qualitative phase of the study undertook in-depth interviews with a subsample of respondents to the survey to explore key quantitative findings, thus providing a deeper and externally valid understanding of the reasons underpinning non-adherence behaviours.

The present study aimed to analyse non-adherence to a cluster of SD rules (keeping 2 mts. distancing, meeting family and friends, and going out for non-essential reasons) using an explanatory sequential mixed-method design. We explored demographic, psycho-social factors and people’s accounts of intentional and unintentional non-adherence to SD rules in a representative sample of North London residents.

## Method

### Design

A mixed-methods, explanatory sequential design (quan→qual) was used, where the qualitative phase (II) used a semi-structured interview guide, informed by key quantitative findings (phase I) obtained in the cross-sectional study [12]. The qualitative phase allowed us to add depth and detail to our associations of variables by further exploring the context, reasons and beliefs underlying non-adherence behaviours.

### Quantitative Phase (I)

#### Participants and Procedure

Over 18s resident in the London boroughs of Islington, Haringey, Camden, Hackney, Barnet or Enfield were surveyed via convenience sampling, using a digital questionnaire delivered through the JISC’s online surveys software, between 1^st^ and 31^st^ May 2020. The total population of the qualifying boroughs is 1,777,666 [13]. In specifying a 99% confidence level and 5% margin of error, the minimum sample size required for this population is 663 [14]. A random prize draw to win one of four £100 vouchers for the Aldi supermarket was used as an incentive to encourage questionnaire completion. The study and the link to the questionnaire were promoted via London Metropolitan University’s website, social media, and local newspapers.

#### Instrument

The digital questionnaire was informed by existing empirical research into factors that have been found to be predictive of protective behaviours during pandemics, as well as two models of health behaviour, the Theory of Planned Behaviour [15] and the Social Ecological Model [10]. Where available, existing scales were used, otherwise items and scales were self-developed. The survey was initially piloted with four academics with expertise in quantitative data and behavioural sciences for expert content validation (conceptual adequacy, relevance, comprehensiveness, and clarity of the items) with slight alterations made subsequently in accordance with the suggestions made. The questionnaire covered the following seven groups of factors:

1. **SD rules infringements.** SD rules infringements were measured via six items, which asked participants to recall SD behaviours from the previous two weeks; how many times participants had gone out for permitted reasons (i.e. for grocery shopping, medication, exercise or work) and not been able to maintain SD (i.e. they came within 2 mts. of someone not lived with); how many times participants broke SD rules to meet up with others (i.e. extended family or friends); and how many times participants went out for unpermitted reasons. All of these counts were totalled to construct the outcome variable of total SD rules infringements. Counts of how many times participants broke SD rules to meet up with others and how many times participants went out for unpermitted reasons were totalled to construct the outcomes variable of intentional SD rules infringements.
2. **Demographic factors.** Demographic data was collected about gender, age, ethnicity, English as a first language, religion, highest qualification obtained, employment status, key worker status and deprivation. Item wording and categories were taken directly from the England Census Rehearsal Household Questionnaire [16]. Deprivation was measured using the English indices of deprivation tool [17], which was determined on the basis of participants’ post codes.
3. **Housing factors.** Participants were asked to identify their housing situation (whether they lived in their own home, a rented home, or a rented room in a house of multiple occupancy), how many people they lived with, and whether they lived with someone vulnerable to COVID-19.
4. **Health factors.** Participants were asked whether, as defined by the UK Government, they had a medical condition which made them more vulnerable to COVID-19 and whether they had experienced COVID-19 symptoms. Perceived susceptibility was measured via a single item, adjusted from a single item measuring perceived susceptibility to cancer [18].
5. **Political factors.** Participants were asked which political party they voted for in the 2019 General Election, which, due to the low number of responses for parties other than Labour or the Conservatives, was recoded as voting for the Conservative government or not. Trust in the Government (3 items, α = .888) was self-developed and covered Government response to COVID-19 and Government follow of scientific advice. Data collection was undertaken during the first national lockdown (1^st^ May to 12^th^ May 2020) and after an easing of restrictions (13^th^ May to 31^st^ May 2020). Participants SD behaviours, recalled over a two-week period, were coded as either being during the total lockdown, during the period of relaxation, or overlapping both periods.
6. **Psychological factors.** COVID-19 and SD knowledge were measured via a self-developed quiz, based upon information from the World Health Organization’s COVID-19 myth busters web portal and from the UK Government’s guidance on SD rules. Self-interest and social responsibility were measured via single items adjusted from Oosterhoff and Palmer [19]. Using the Theory of Planned Behaviour as a guide, SD behavioural intention (3 items, α = .854) was self-developed. Three perceived behavioural control items were self-developed measuring control over leaving the house, control over others’ distancing and control over responsibilities. Normative pressure from family, friends and neighbours were each measured via self-developed single items.
7. **Social factors.** Based upon the Social Ecological Model, participants were asked to report if they were receiving financial and community support if needed during lockdown. Social support was measured using the multidimensional scale of perceived social support [20], with items contextualised to refer to the lockdown period. Sub-scales for support from a special person (3 items, α = .939), family (3 items, α = .937) and friends (3 items, α = .94) were used.

#### Statistical analysis

To measure the associations between explanatory variables (i.e. demographic, housing, health, political, psychological and social factors) and outcome variables (i.e. total infringements and intentional infringements of SD rules) univariate and multivariate analysis were undertaken, so to provide a complete picture of explanatory factors, including those that are significant in univariate analysis but which are not significant in multivariate analysis when other factors better accounted for variance. For univariate analysis, independent samples t-tests and one-way ANOVA tests were ran to identify statistically significant differences in mean infringements between categories of categorical explanatory variables. Also, Pearson’s product-moment (for interval and ratio scale explanatory variables) and Spearman’s rank order (for ordinal ratio scale variables) correlations were ran to identify statistically significant associations between numerical explanatory variables and infringements.

The outcome variables of total infringements and intentional infringements are count data, as such a Poisson regression model was used in the first instance. An assumption of this model is that the distribution of counts follows a Poisson distribution, such that the observed and expected counts should be approximately equal. However, for both models, a Pearson Chi-Square goodness of fit test returned a value considerably greater than 1 (total infringements χ^2^ (589) = 6.631; intentional infringements χ^2^ (589) = 6.247), indicating overdispersion whereby the observed total infringements and intentional infringements exceeded the expected total infringements and intentional infringements. Given that that the assumption of equidispersion was not met, a negative binomial regression model was used instead. This is another model for count data, which allows for overdispersion because it can take on a more varied set of shapes than the Poisson distribution, such that the assumptions for this model are met by the data.

### Qualitative Phase (II)

#### Participants and recruitment

For the qualitative study, we used the quantitative findings to inform our sampling plan. We developed a matrix to purposively sampling relevant socio-demographic groups based on age, gender, ethnicity, employment status, borough, and clinical vulnerability. After stratification, we conducted a random selection from the survey data amongst those individuals who consented to be contacted for an interview. Individuals were invited by email to an interview to be conducted by phone or online platforms, Zoom or Skype, according to their preference, and were offered a £20 Aldi voucher for their participation.

#### Data collection

Interviews were conducted by phone (n=9), Zoom and Skype (n=21) between the 5^th^ August and 21st September 2020. Interviews were conducted by YE, an experienced qualitative researcher with a background in health studies and public health, who emphasised to participants the study was non-judgemental about their level of adherence to SD measures, and we were only interested in the reasons and motivations for their behaviours. Interviewees were asked to confirm demographic information collected in the survey, and any changes observed (e.g. employment status or housing situation) were recorded.

We used semi-structured in-depth interviews with a topic guide aimed to further interpret and explain our quantitative results, including counterintuitive findings. The interview guide comprised of open-ended questions, each with several prompts. It was reviewed by key stakeholders (see public involvement below). The guide included questions at the individual, interpersonal and community levels following the Social Ecological Model approach, including the Theory of Planned Behaviour for individual factors. Individual level factors covered were perceived behavioural control [self-efficacy and controllability], social norms [friends, family, and neighbours], perceived threat [vulnerability, susceptibility and severity], attitude towards norms and trust in government, and intentions). Interpersonal level factors included social support [friends, family, and statutory services]. Community level factors covered were local and environmental area perceptions.

All interviews were digitally recorded and transcribed verbatim. Participants have been anonymised, given identifier codes, and age ranges. For example, F02, WEG, 35+ means that interviewee is a female, interview number 02, White ethnic group, age range 35-39 years old. Age range such as ‘40s’ means [40-44 years old]; ‘O’ next to a number (gender other); ‘OEG’ (Other ethnic group); ‘BEG’ (Black ethnic group); ‘AEG’ (Asian ethnic group).

#### Public involvement

The interview guide was reviewed by key stakeholders from local Public Health, Healthwatch and NHS Northcentral London CCG with whom quantitative findings were discussed during a workshop. Feedback was incorporated into the interview guide before recruitment started. Due to the tight schedule of this research, it was not possible to involve the general public in the development of the interview or survey instruments.

#### Analysis

Framework analysis [21] was used for qualitative data interpretation, which involves five stages (familiarization with the data, identification of a thematic framework, indexing, charting, and mapping and interpretation of themes). YE read all the transcripts and both YE and SH read a selection of transcripts to identify recurring themes. Both researchers independently coded the same five transcripts that led to the development of a coding framework. Coded transcripts were compared, and codes further refined and grouped into broader categories. Categories were derived deductively based on the Social Ecological Model and codes were derived both deductively and inductively based on the Theory of Planned Behaviour, quantitative findings, and participants’ accounts. The complete dataset was then manually indexed by YE, and SH independently coded two interviews to ensure consistency. Once indexing was complete, data was arranged in a case chart with one row per participants and one category and associated codes per column, alongside illustrative quotes. This allowed for further identification of patterns and associations (i.e. similarities and differences in relation to participants’ age, gender and ethnicity) during the mapping and interpretation process, which led to the generation of themes across the case chart and the research questions.

### Data integration

In this paper, we merged results from both sets of data analyses known as ‘mixing during interpretation’ in the Discussion section [12]. In the process of integration, we aimed for ‘expansion’ as qualitative data had the purpose of explaining the nature of the associations observed in the quantitative data [22].

### Ethics

Ethical approval for this study was granted by London Metropolitan University Research Ethics Committee (reference: GSBL200401).

A participant information sheet (PIS) was provided, and informed consent (IC) was obtained from all participants before completion of the online survey; and for qualitative data collection PIS and IC were provided by email with consent forms orally recorded during the interviews or retuned by email.

## Quantitative Results

### Participants

There were a total of 681 valid responses to the study’s questionnaire. There were 20 responses from participants living in locations other than the specified North London boroughs, which were removed from the dataset. The characteristics of the final sample are reported in Table 2. Of note is that the sample was highly skewed to females, with 82.8% of respondents being female (564 vs. 111 males), which is reflective of the trend that women are more likely to participate in surveys than men [23–24]. Also of note is that a minority of 14.4% of participants came from BAME populations (98 vs. 583 White), which is not reflective of the broader London population, 40.2% of whom come from BAME groups [25]. This under-representation reflects the well-established trend that ethnic minorities are less likely to participate in health surveys than ethnic majorities [26–27].

**Table 1.**
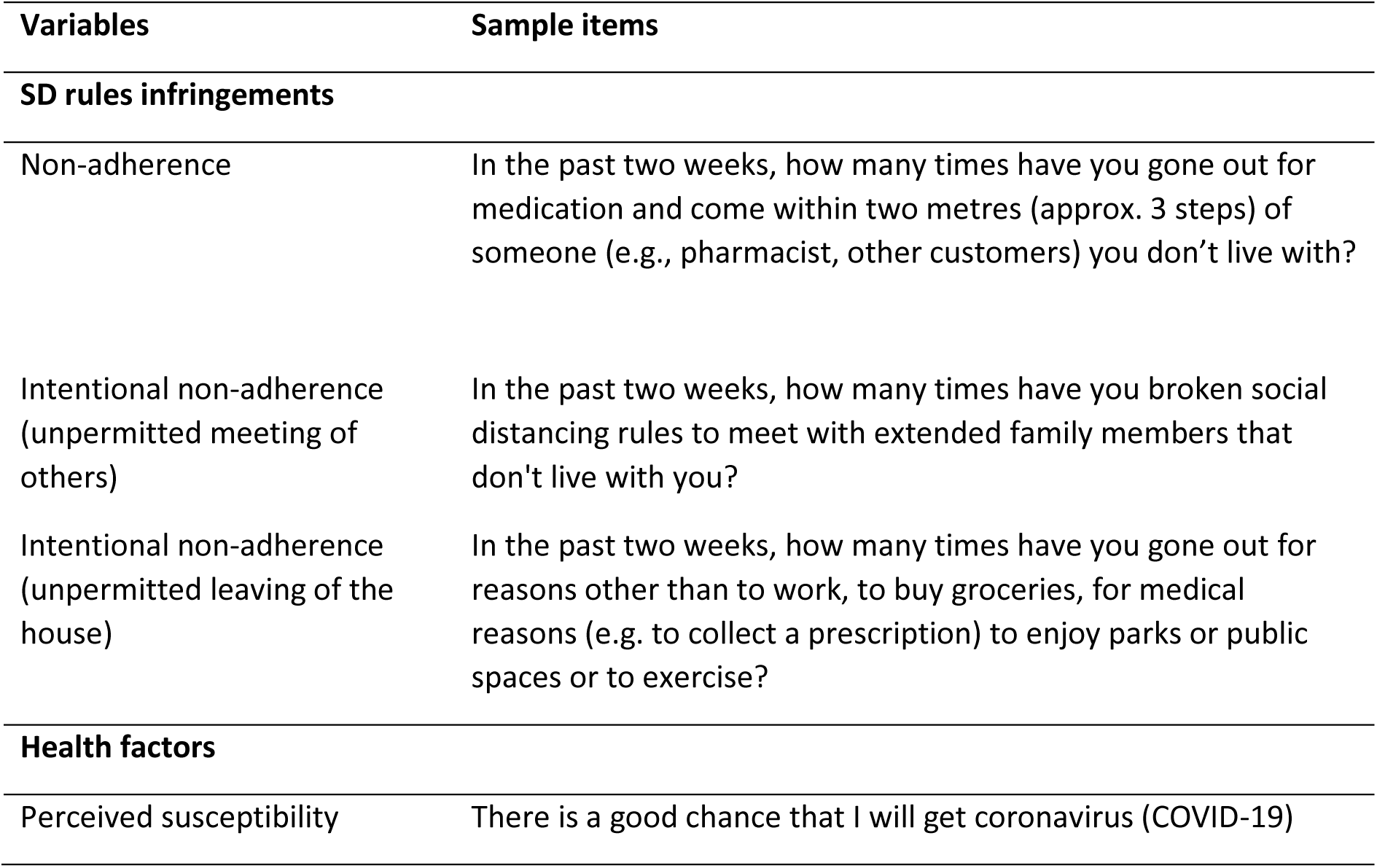
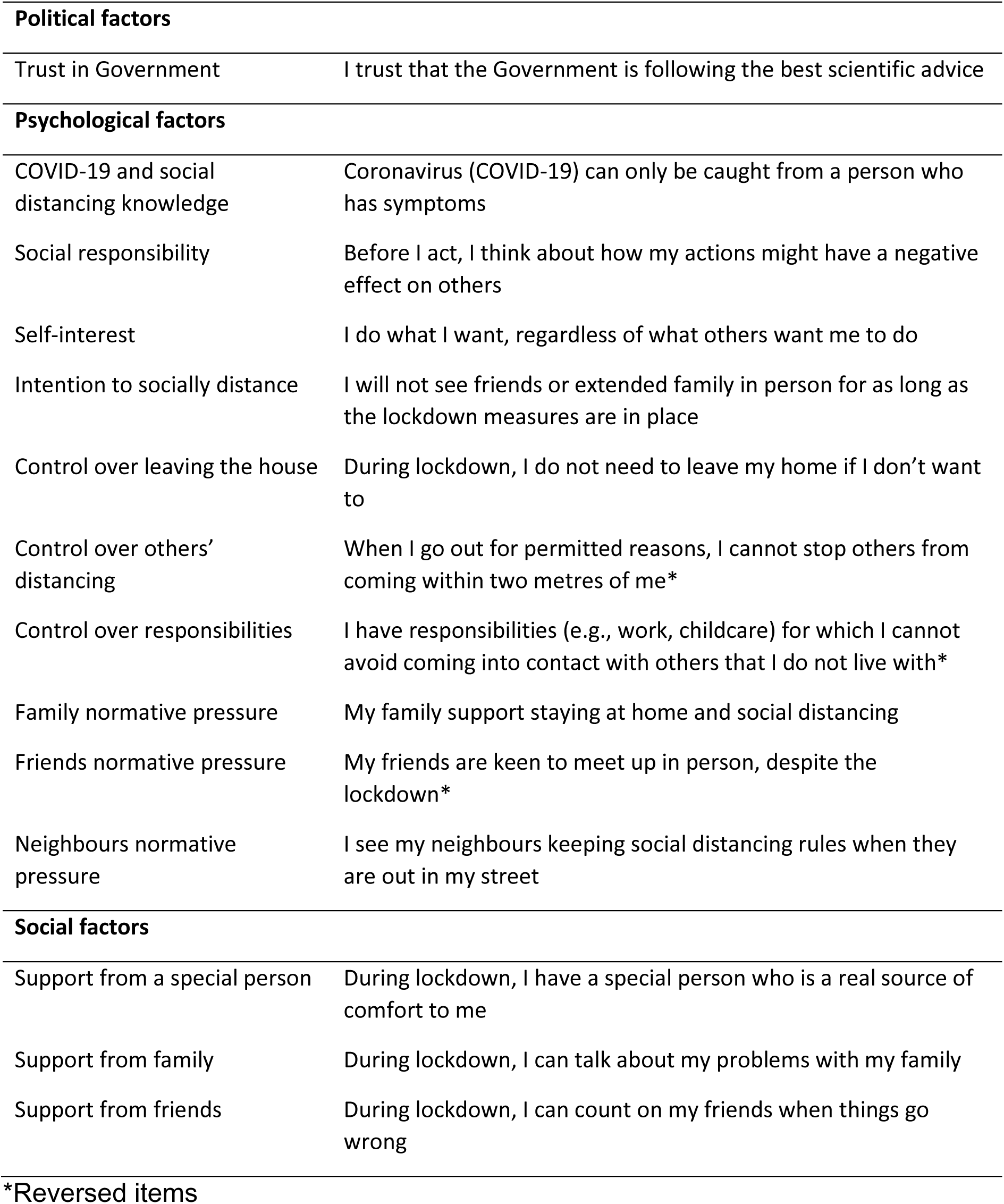
Sample items of research variables

| Variables | Sample items |
| --- | --- |
| **SD rules infringements** |  |
| Non-adherence | In the past two weeks, how many times have you gone out for medication and come within two metres (approx. 3 steps) of someone (e.g., pharmacist, other customers) you don't live with? |
| Intentional non-adherence (unpermitted meeting of others) | In the past two weeks, how many times have you broken social distancing rules to meet with extended family members that don't live with you? |
| Intentional non-adherence (unpermitted leaving of the house) | In the past two weeks, how many times have you gone out for reasons other than to work, to buy groceries, for medical reasons (e.g. to collect a prescription) to enjoy parks or public spaces or to exercise? |
| **Health factors** |  |
| Perceived susceptibility | There is a good chance that I will get coronavirus (COVID-19) |
| **Political factors** |  |
| Trust in Government | I trust that the Government is following the best scientific advice |
| **Psychological factors** |  |
| COVID-19 and social distancing knowledge | Coronavirus (COVID-19) can only be caught from a person who has symptoms |
| Social responsibility | Before I act, I think about how my actions might have a negative effect on others |
| Self-interest | I do what I want, regardless of what others want me to do |
| Intention to socially distance | I will not see friends or extended family in person for as long as the lockdown measures are in place |
| Control over leaving the house | During lockdown, I do not need to leave my home if I don't want to |
| Control over others' distancing | When I go out for permitted reasons, I cannot stop others from coming within two metres of me* |
| Control over responsibilities | I have responsibilities (e.g., work, childcare) for which I cannot avoid coming into contact with others that I do not live with* |
| Family normative pressure | My family support staying at home and social distancing |
| Friends normative pressure | My friends are keen to meet up in person, despite the lockdown* |
| Neighbours normative pressure | I see my neighbours keeping social distancing rules when they are out in my street |
| **Social factors** |  |
| Support from a special person | During lockdown, I have a special person who is a real source of comfort to me |
| Support from family | During lockdown, I can talk about my problems with my family |
| Support from friends | During lockdown, I can count on my friends when things go wrong |

\*Reversed items

**Table 2.**
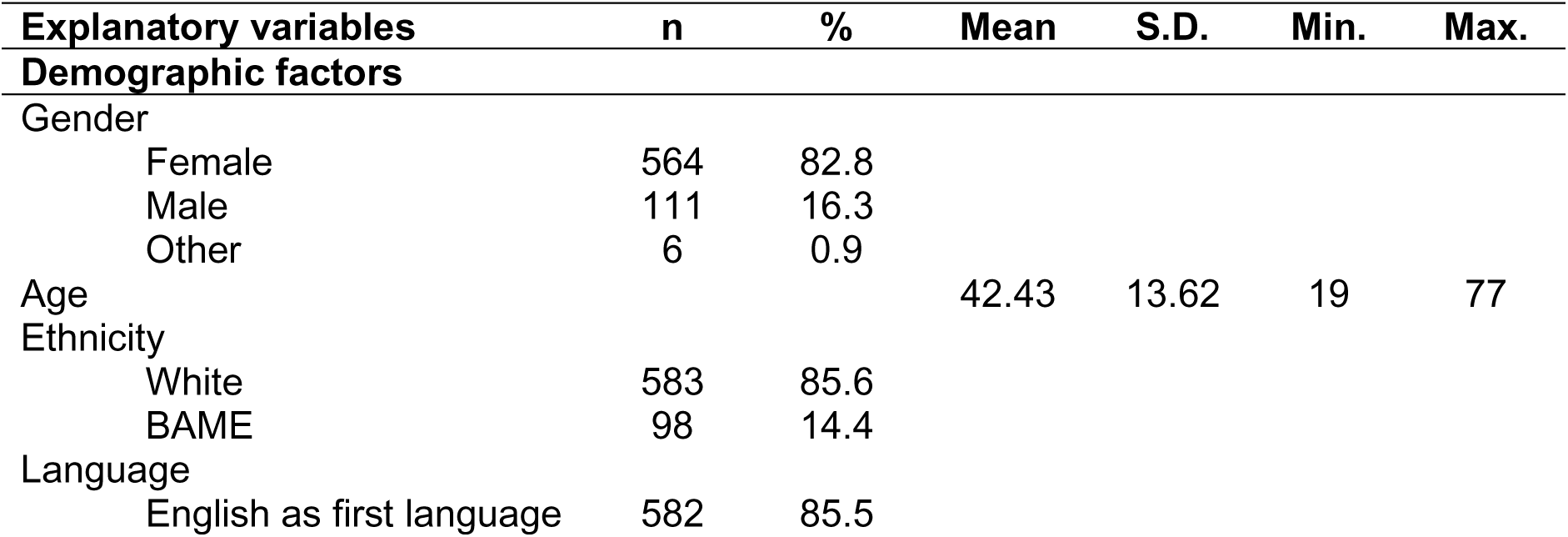
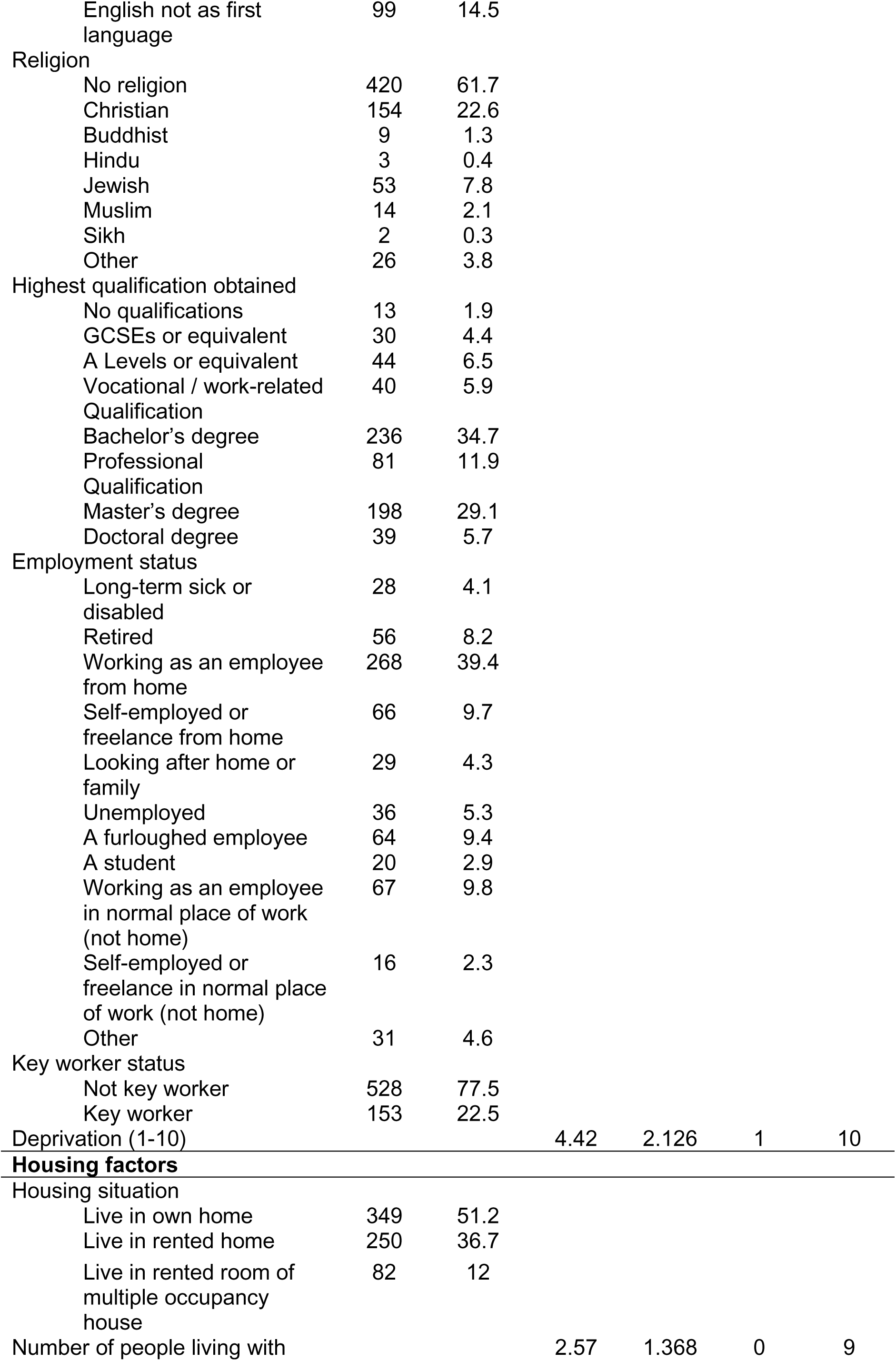
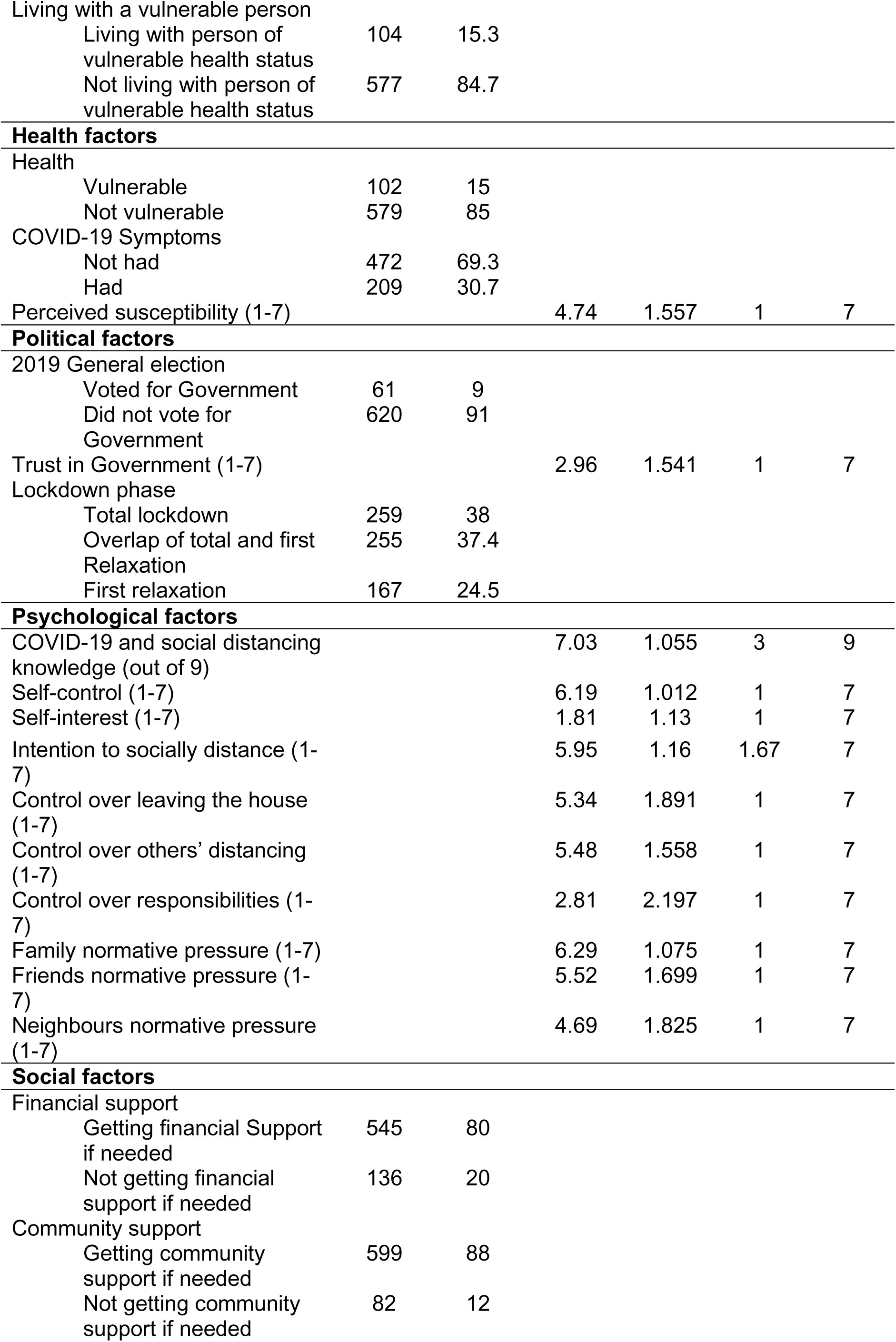
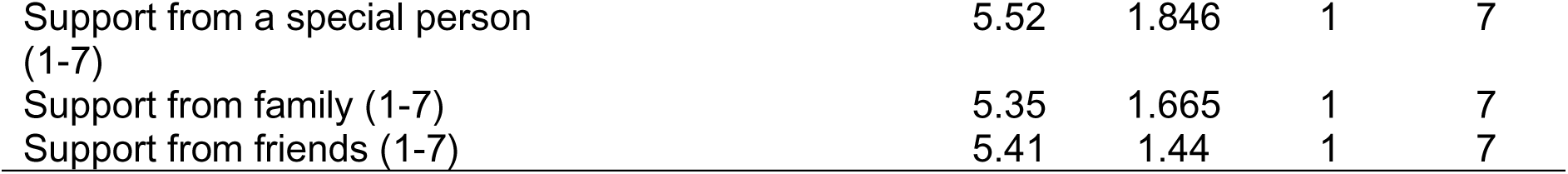
Characteristics of sample

| Explanatory variables | n | % | Mean | S.D. | Min. | Max. |
| --- | --- | --- | --- | --- | --- | --- |
| **Demographic factors** |  |  |  |  |  |  |
| Gender |  |  |  |  |  |  |
| Female | 564 | 82.8 |  |  |  |  |
| Male | 111 | 16.3 |  |  |  |  |
| Other | 6 | 0.9 |  |  |  |  |
| Age |  |  | 42.43 | 13.62 | 19 | 77 |
| Ethnicity |  |  |  |  |  |  |
| White | 583 | 85.6 |  |  |  |  |
| BAME | 98 | 14.4 |  |  |  |  |
| Language |  |  |  |  |  |  |
| English as first language | 582 | 85.5 |  |  |  |  |
| English not as first language | 99 | 14.5 |  |  |  |  |
| Religion |  |  |  |  |  |  |
| No religion | 420 | 61.7 |  |  |  |  |
| Christian | 154 | 22.6 |  |  |  |  |
| Buddhist | 9 | 1.3 |  |  |  |  |
| Hindu | 3 | 0.4 |  |  |  |  |
| Jewish | 53 | 7.8 |  |  |  |  |
| Muslim | 14 | 2.1 |  |  |  |  |
| Sikh | 2 | 0.3 |  |  |  |  |
| Other | 26 | 3.8 |  |  |  |  |
| Highest qualification obtained |  |  |  |  |  |  |
| No qualifications | 13 | 1.9 |  |  |  |  |
| GCSEs or equivalent | 30 | 4.4 |  |  |  |  |
| A Levels or equivalent | 44 | 6.5 |  |  |  |  |
| Vocational / work-related Qualification | 40 | 5.9 |  |  |  |  |
| Bachelor's degree | 236 | 34.7 |  |  |  |  |
| Professional Qualification | 81 | 11.9 |  |  |  |  |
| Master's degree | 198 | 29.1 |  |  |  |  |
| Doctoral degree | 39 | 5.7 |  |  |  |  |
| Employment status |  |  |  |  |  |  |
| Long-term sick or disabled | 28 | 4.1 |  |  |  |  |
| Retired | 56 | 8.2 |  |  |  |  |
| Working as an employee from home | 268 | 39.4 |  |  |  |  |
| Self-employed or freelance from home | 66 | 9.7 |  |  |  |  |
| Looking after home or family | 29 | 4.3 |  |  |  |  |
| Unemployed | 36 | 5.3 |  |  |  |  |
| A furloughed employee | 64 | 9.4 |  |  |  |  |
| A student | 20 | 2.9 |  |  |  |  |
| Working as an employee in normal place of work (not home) | 67 | 9.8 |  |  |  |  |
| Self-employed or freelance in normal place of work (not home) | 16 | 2.3 |  |  |  |  |
| Other | 31 | 4.6 |  |  |  |  |
| Key worker status |  |  |  |  |  |  |
| Not key worker | 528 | 77.5 |  |  |  |  |
| Key worker | 153 | 22.5 |  |  |  |  |
| Deprivation (1-10) |  |  | 4.42 | 2.126 | 1 | 10 |
| **Housing factors** |  |  |  |  |  |  |
| Housing situation |  |  |  |  |  |  |
| Live in own home | 349 | 51.2 |  |  |  |  |
| Live in rented home | 250 | 36.7 |  |  |  |  |
| Live in rented room of multiple occupancy house | 82 | 12 |  |  |  |  |
| Number of people living with |  |  | 2.57 | 1.368 | 0 | 9 |
| Living with a vulnerable person |  |  |  |  |  |  |
| Living with person of vulnerable health status | 104 | 15.3 |  |  |  |  |
| Not living with person of vulnerable health status | 577 | 84.7 |  |  |  |  |
| **Health factors** |  |  |  |  |  |  |
| Health |  |  |  |  |  |  |
| Vulnerable | 102 | 15 |  |  |  |  |
| Not vulnerable | 579 | 85 |  |  |  |  |
| COVID-19 Symptoms |  |  |  |  |  |  |
| Not had | 472 | 69.3 |  |  |  |  |
| Had | 209 | 30.7 |  |  |  |  |
| Perceived susceptibility (1-7) |  |  | 4.74 | 1.557 | 1 | 7 |
| **Political factors** |  |  |  |  |  |  |
| 2019 General election |  |  |  |  |  |  |
| Voted for Government | 61 | 9 |  |  |  |  |
| Did not vote for Government | 620 | 91 |  |  |  |  |
| Trust in Government (1-7) |  |  | 2.96 | 1.541 | 1 | 7 |
| Lockdown phase |  |  |  |  |  |  |
| Total lockdown | 259 | 38 |  |  |  |  |
| Overlap of total and first Relaxation | 255 | 37.4 |  |  |  |  |
| First relaxation | 167 | 24.5 |  |  |  |  |
| **Psychological factors** |  |  |  |  |  |  |
| COVID-19 and social distancing knowledge (out of 9) |  |  | 7.03 | 1.055 | 3 | 9 |
| Self-control (1-7) |  |  | 6.19 | 1.012 | 1 | 7 |
| Self-interest (1-7) |  |  | 1.81 | 1.13 | 1 | 7 |
| Intention to socially distance (1-7) |  |  | 5.95 | 1.16 | 1.67 | 7 |
| Control over leaving the house (1-7) |  |  | 5.34 | 1.891 | 1 | 7 |
| Control over others' distancing (1-7) |  |  | 5.48 | 1.558 | 1 | 7 |
| Control over responsibilities (1-7) |  |  | 2.81 | 2.197 | 1 | 7 |
| Family normative pressure (1-7) |  |  | 6.29 | 1.075 | 1 | 7 |
| Friends normative pressure (1-7) |  |  | 5.52 | 1.699 | 1 | 7 |
| Neighbours normative pressure (1-7) |  |  | 4.69 | 1.825 | 1 | 7 |
| **Social factors** |  |  |  |  |  |  |
| Financial support |  |  |  |  |  |  |
| Getting financial Support if needed | 545 | 80 |  |  |  |  |
| Not getting financial support if needed | 136 | 20 |  |  |  |  |
| Community support |  |  |  |  |  |  |
| Getting community support if needed | 599 | 88 |  |  |  |  |
| Not getting community support if needed | 82 | 12 |  |  |  |  |

|  |  |  |  |  |
| --- | --- | --- | --- | --- |
| Support from a special person (1-7) | 5.52 | 1.846 | 1 | 7 |
| Support from family (1-7) | 5.35 | 1.665 | 1 | 7 |
| Support from friends (1-7) | 5.41 | 1.44 | 1 | 7 |

### SD rules infringements

As reported in Table 3, over a period of two weeks, the average number of infringements of any kind per study participant was 10.77. Of these, on average, participants unintentionally broke SD rules (i.e. unable to maintain social distance from others when going out for permitted reasons) 8.18 times. Out of 10.77 total infringements, participants, on average, intentionally broke the rules (i.e. left house for unpermitted reasons or engaged in non-permitted meeting of others) 2.59 times. Of these, participants, on average, left their house for unpermitted reasons 1.92 times and engaged in unpermitted meeting of others 0.67 times.

**Table 3.** Infringements of SD rules

|  | Mean | Std. Deviation | Minimum | Maximum |
| --- | --- | --- | --- | --- |
| All infringements | 10.77 | 9.36 | 0 | 59 |
| Unintentional infringements | 8.18 | 7.26 | 0 | 42 |
| Intentional infringements | 2.59 | 4.62 | 0 | 32 |
| Unpermitted leaving of house infringements | 1.92 | 4.07 | 0 | 30 |
| Unpermitted meeting of others infringements | 0.67 | 1.62 | 0 | 20 |

### Factors associated with SD infringements

#### Univariate analysis

The differences in means of infringements between categories of each categorical explanatory variable are reported in Table 4. There was a statistically significant difference in infringements between housing situation groups as determined by a one-way ANOVA (F(2,678) = 3.59, p= .028). A LSD post hoc test revealed that infringements were statistically significantly higher for participants living in a rented home (11.64 ± 9.56) compared to participants who live in their own home (9.85 ± 9.25, p = .02). An independent-samples t-test found that participants who were not vulnerable committed statistically significantly more infringements (11.38 ± 9.32) compared to participants who were vulnerable (7.29 ± 8.84), a mean difference of 4.09 (95% CI, 2.14 to 6.04), t(679) = 4.12, p = .000.

**Table 4.** Average infringements by category of categorical explanatory variable

| Explanatory variables | All infringements |  | Intentional infringements |  |
| --- | --- | --- | --- | --- |
|  | Mean | S.D. | Mean | S.D. |
| Sample | 10.77 | 9.36 | 2.59 | 4.62 |
| **Demographic factors** |  |  |  |  |
| Gender |  |  |  |  |
| Female | 10.73 | 9.01 | 2.76 | 5 |
| Male | 11.27 | 9.45 | 2.57 | 4.56 |
| Other | 5 | 5.33 | 1.5 | 2.74 |
| Ethnicity |  |  |  |  |
| White | 10.9 | 9.4 | 2.64 | 4.63 |
| BAME | 10.01 | 9.16 | 2.33 | 4.59 |
| Language |  |  |  |  |
| English as first language | 10.78 | 9.56 | 2.6 | 4.75 |
| English not as first language | 10.73 | 8.17 | 2.55 | 3.83 |
| Religion |  |  |  |  |
| No religion | 10.65 | 9.02 | 2.52 | 4.38 |
| Christian | 11.34 | 10.06 | 2.94 | 5.17 |
| Buddhist | 10 | 8.09 | 3.67 | 5.36 |
| Hindu | 4.33 | 3.79 | .67 | 1.16 |
| Jewish | 11.38 | 10.63 | 3.23 | 5.8 |
| Muslim | 8.57 | 7.92 | 2.07 | 3.63 |
| Sikh | 9.5 | 9.19 | 0 | .00 |
| Other | 10.38 | 9.78 | .85 | 1.74 |
| Highest qualification obtained |  |  |  |  |
| No qualifications | 16.62 | 14.33 | 3.92 | 5.38 |
| GCSEs or equivalent | 10.9 | 7.88 | 3.73 | 5.17 |
| A Levels or equivalent | 8.8 | 7.25 | 2.27 | 3.99 |
| Vocational / work-related qualification | 9.05 | 9.17 | 1.55 | 2.86 |
| Bachelor's degree | 11.16 | 10.17 | 2.69 | 4.66 |
| Professional qualification | 9.37 | 7.81 | 2 | 3.66 |
| Master's degree | 10.84 | 8.88 | 2.68 | 5.17 |
| Doctoral degree | 12.95 | 10.21 | 2.87 | 4.6 |
| Employment Status |  |  |  |  |
| Long-term sick or disabled | 8.86 | 10.24 | 1.32 | 2.76 |
| Retired | 7.88 | 10.4 | 3.09 | 6.36 |
| Working as an employee from home | 10.85 | 9.64 | 2.31 | 4.58 |
| Self-employed or freelance from home | 10.39 | 8.04 | 2.03 | 4 |
| Looking after home or family | 9.59 | 6.76 | 2.97 | 4.15 |
| Unemployed | 10.17 | 8.29 | 2.11 | 3.76 |
| A furloughed employee | 12.97 | 10.19 | 3.69* | 5.07 |
| A student | 11.95 | 8.78 | 3.85 | 4.67 |
| Working as an employee in normal place of work (not home) | 12.03 | 8.5 | 2.37 | 4.07 |
| Self-employed or freelance in normal place of work (not home) | 15.06 | 9.26 | 5.81* | 5.28 |
| Other | 9.39 | 9.34 | 2.48 | 4.15 |
| Key Worker status |  |  |  |  |
| Not key worker | 10.57 | 9.31 | 2.54 | 4.55 |
| Key worker | 11.45 | 9.53 | 2.79 | 4.64 |
| **Housing factors** |  |  |  |  |
| Housing situation |  |  |  |  |
| Live in own home | 9.85 | 9.25 | 2.67 | 4.92 |
| Live in rented home | 11.64* | 9.56 | 2.47 | 4.39 |
| Live in rented room of multiple occupancy house | 12.06 | 8.94 | 2.66 | 3.99 |
| Living with a vulnerable person |  |  |  |  |
| Living with person of vulnerable health status | 9.36 | 9.1 | 2.25 | 4.64 |
| Not living with person of vulnerable health status | 11.03 | 9.39 | 2.66 | 4.62 |
| **Health factors** |  |  |  |  |
| Vulnerable health |  |  |  |  |
| Vulnerable | 7.29 | 8.84 | 1.75 | 4.16 |
| Not vulnerable | 11.38* | 9.32 | 2.74* | 4.68 |
| COVID-19 Symptoms |  |  |  |  |
| Not had | 10.6 | 9.53 | 2.52 | 4.73 |
| Had | 11.17 | 8.97 | 2.76 | 4.37 |
| **Political factors** |  |  |  |  |
| 2019 General election |  |  |  |  |
| Voted for Government | 11 | 10.65 | 3.7 | 5.83 |
| Did not vote for Government | 10.75 | 9.23 | 2.48 | 4.48 |
| Lockdown phase |  |  |  |  |
| Total lockdown | 10.47 | 9.21 | 2.08 | 4.12 |
| Overlap of total and first relaxation | 11.12 | 9.07 | 2.58 | 4.36 |
| First relaxation | 10.71 | 10.06 | 3.4* | 5.56 |
| **Social factors** |  |  |  |  |
| Financial support |  |  |  |  |
| Getting financial support if needed | 10.91 | 9.53 | 2.68 | 4.85 |
| Not getting financial support if needed | 10.21 | 8.68 | 2.26 | 3.56 |
| Community support |  |  |  |  |
| Getting community support if needed | 10.88 | 9.46 | 2.64 | 4.72 |
| Not getting community support if needed | 9.96 | 8.58 | 2.23 | 3.8 |

\*Statistically significantly higher infringements

The associations between numerical explanatory variables and infringements are reported in Table 5. There was a moderate, negative correlation between intention to socially distance and infringements (*r_s_*(681) = -.337, *p* = .000). There was a weak, negative correlation between age (*r* (681) = -.104, *p* = .006), social responsibility (*r_s_*(681) = -.097, *p* = .012) control over leaving the house (*r_s_*(681) = -.208, *p* = .000), control over others’ distancing (*r_s_*(681) = -.217, *p* = .000) control over responsibilities (*r_s_*(681) = -.204, *p* = .000), normative pressure from family (*r_s_*(681) = -.117, *p* = .002), normative pressure from friends and infringements (*r_s_*(681) = -.222, *p* = .000) and total infringements. There was a weak, positive correlation between support from friends and infringements (*r_s_*(681) = .097, *p* = .011).

**Table 5.** Associations between numerical explanatory variables and infringements

| Explanatory variables | All infringements |  | Intentional infringements |  |
| --- | --- | --- | --- | --- |
|  | Correlation | Sig. | Correlation | Sig. |
| **Demographic factors** |  |  |  |  |
| Age | -.104* | .006 | .053 | .169 |
| Deprivation | -.045 | .243 | .015 | .702 |
| **Housing factor** |  |  |  |  |
| Number of people living with | .047 | .216 | .044 | .257 |
| **Health factor** |  |  |  |  |
| Perceived susceptibility | .067 | .079 | .061 | .114 |
| **Political factor** |  |  |  |  |
| Trust in Government | -.034 | .377 | -.033 | .389 |
| **Psychological factors** |  |  |  |  |
| COVID-19 and social distancing knowledge | -.062 | .106 | -.08* | .036 |
| Social responsibility | -.097* | .012 | -.147* | .000 |
| Self-interest | .057 | .141 | .143* | .000 |
| Intention to socially distance | -.337* | .000 | -.409* | .000 |
| Control over leaving the house | -.208* | .000 | -.1* | .009 |
| Control over others' distancing | -.217* | .000 | -.09* | .019 |
| Control over responsibilities | -.204* | .000 | -.13* | .001 |
| Normative pressure from family | -.117* | .002 | -.189* | .000 |
| Normative pressure from friends | -.222* | .000 | -.21* | .000 |
| Normative pressure from neighbours | -.011 | .768 | .044 | .247 |
| **Social Factors** |  |  |  |  |
| Support from a special person | .032 | .411 | .001 | .978 |
| Support from family | .023 | .554 | -.001 | .987 |
| Support from friends | .097* | .011 | .06 | .117 |

\*Statistically significant univariate association

**Table 6.**
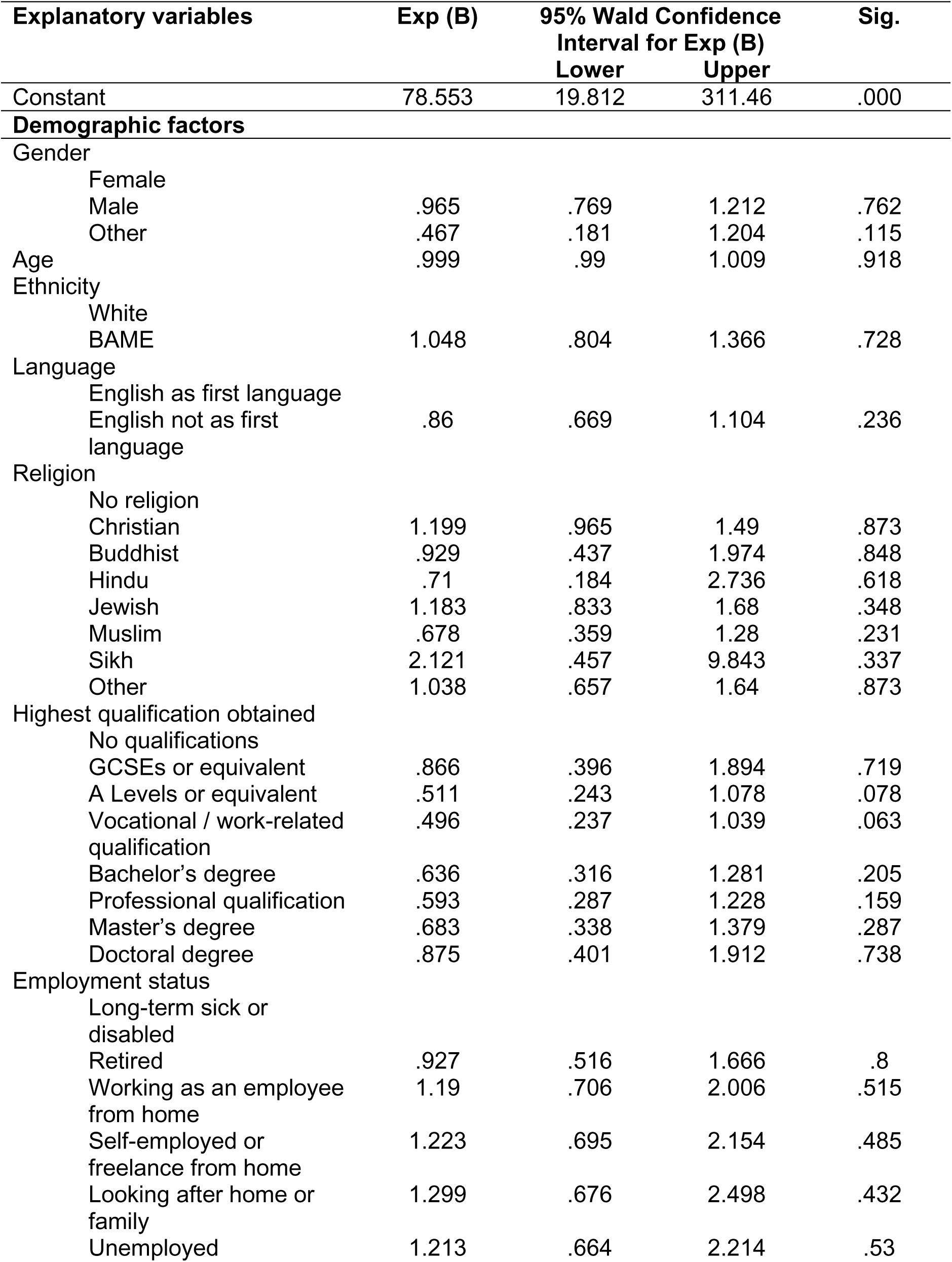
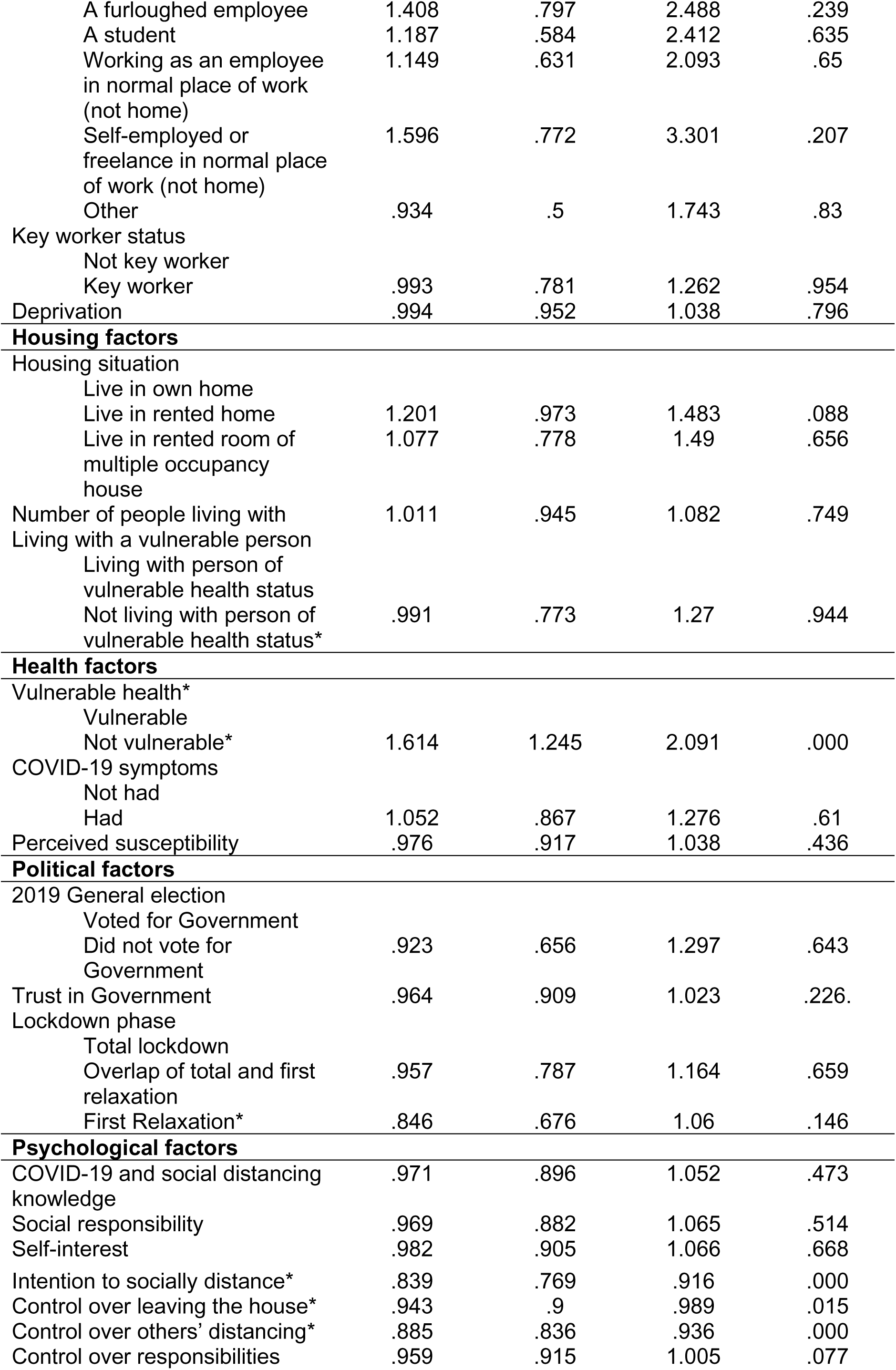
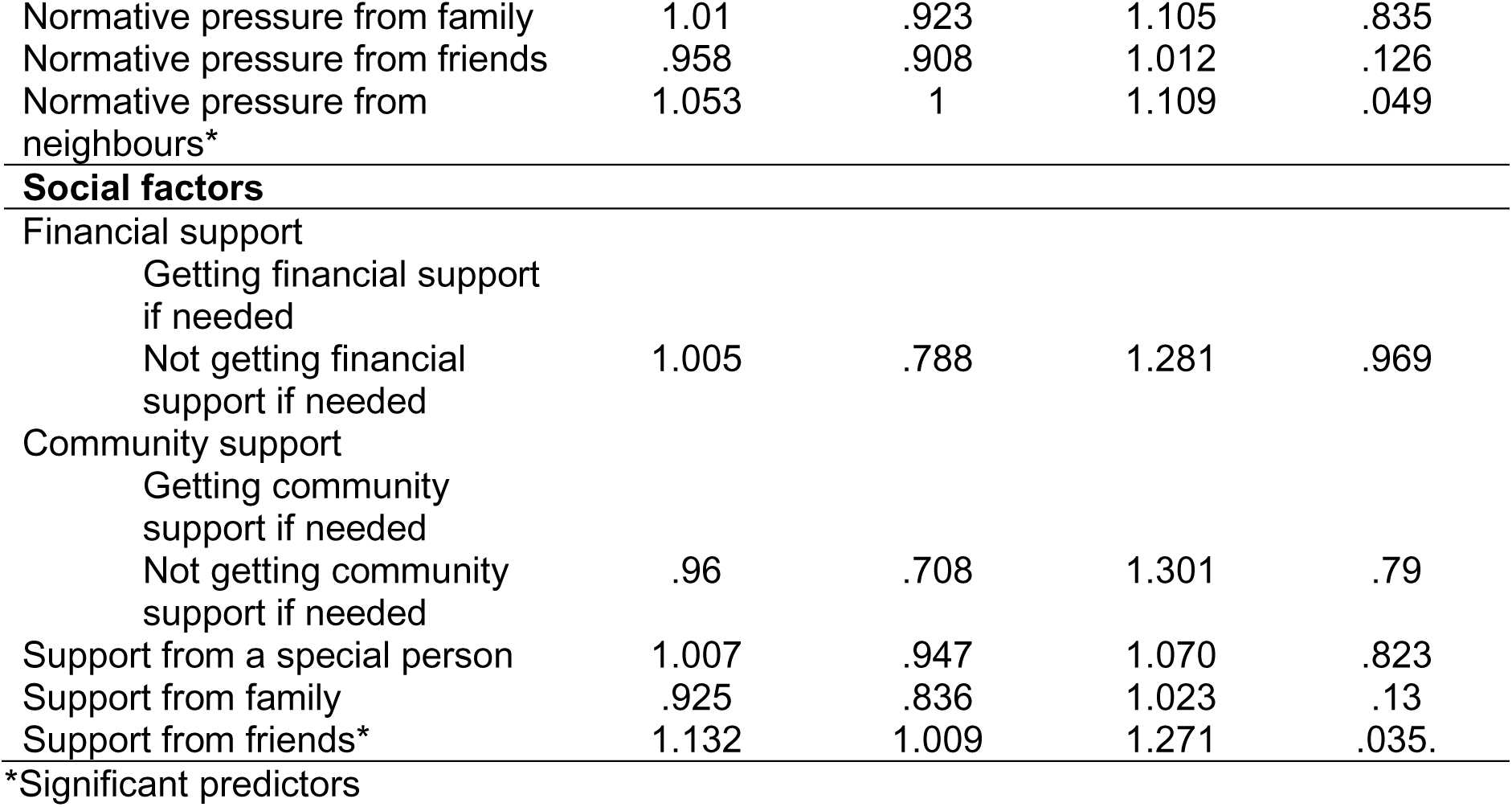
Results of negative binomial regression, with count outcome variable of total infringements of social distancing rules

**Table 7.**
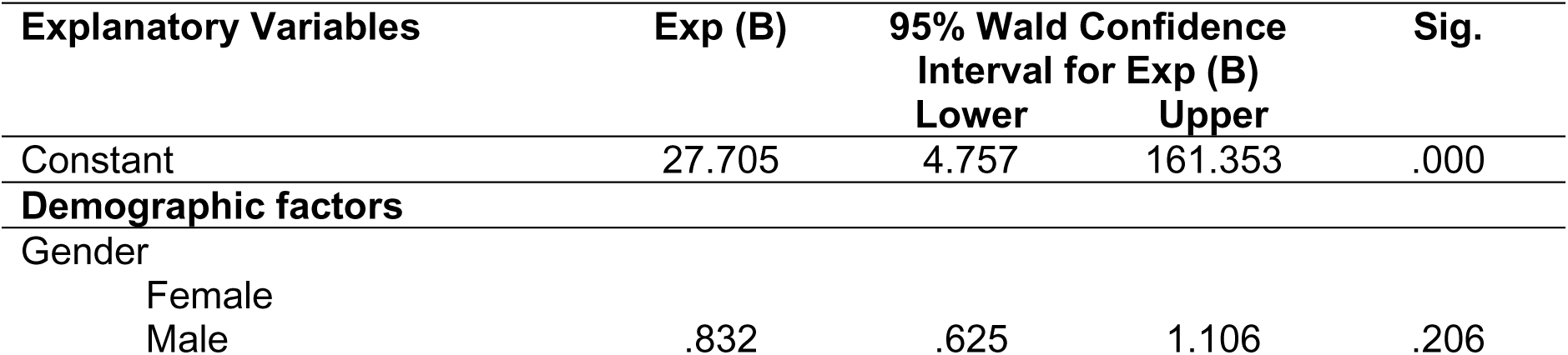
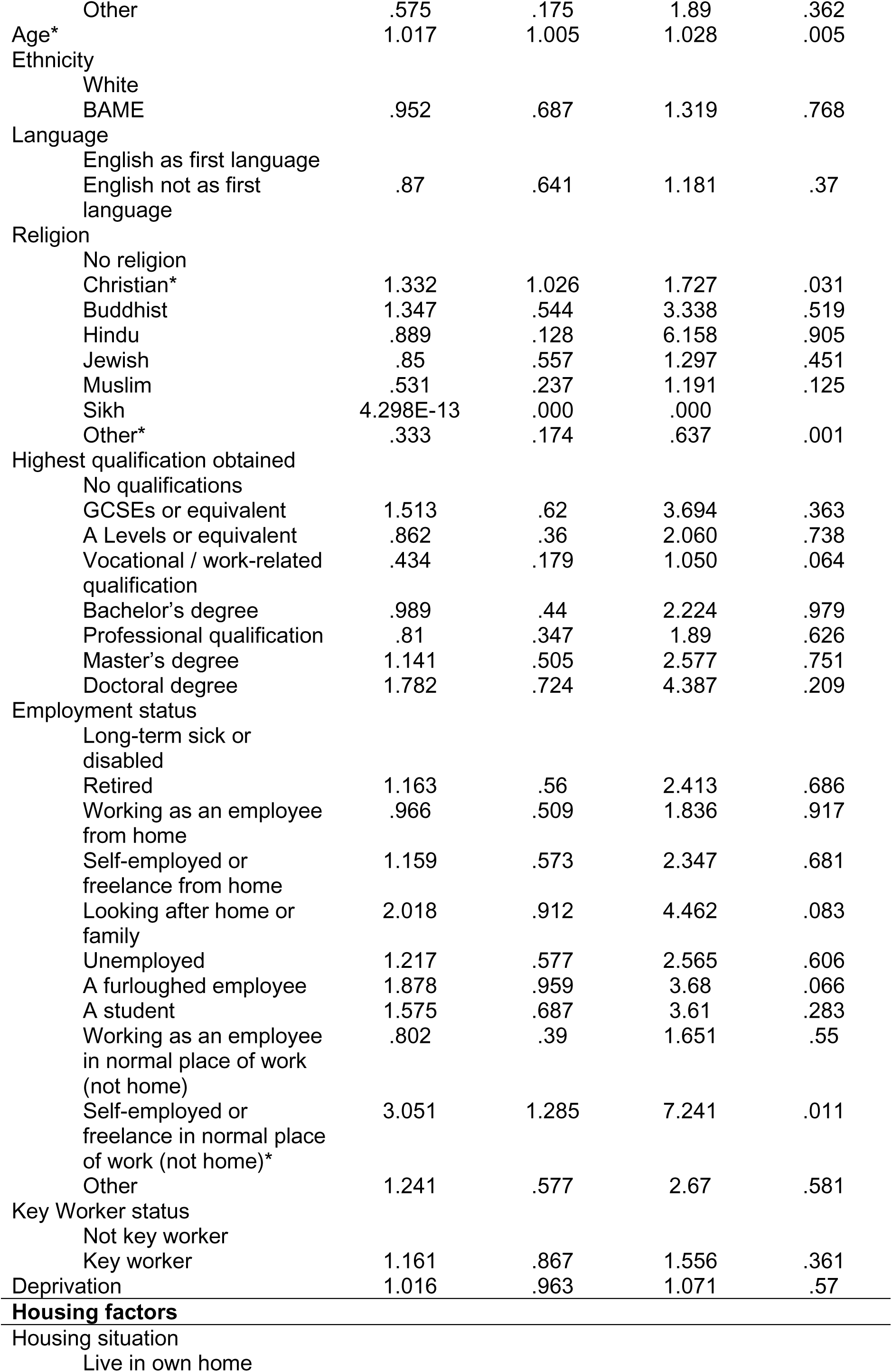
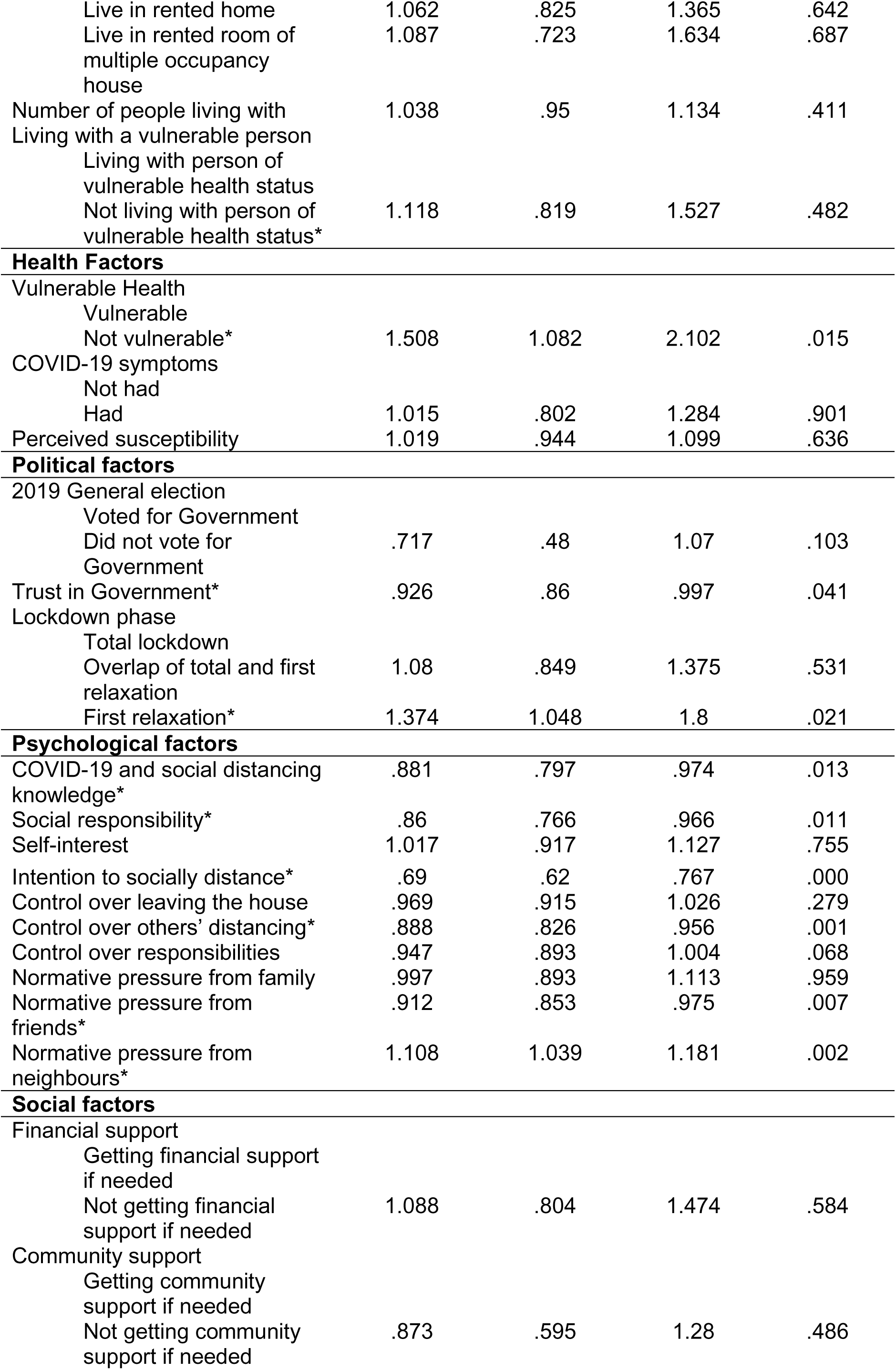
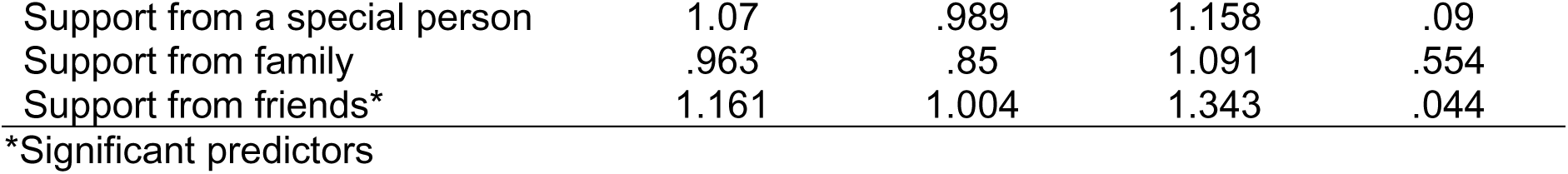
Results of negative binomial regression, with count outcome variable of intentional infringements of social distancing rules

#### Multivariate analysis

The negative binomial regression model was statistically significant, χ^2^(57) = 129.131, *p* = .000. A Pearson Chi-Square goodness of fit test returned a value greater than 0.5 (χ^2^ (623) = .628), indicating that the model fits the data well. When holding other factors constant, SD infringements increase by 61.4% if an individual is not vulnerable, than if an individual is vulnerable. An additional level of agreement on a 7-point Likert scale indicating intention to socially distance decreases SD infringements by 16.1%. An additional level of agreement on a 7-point Likert scale indicating having control over leaving the house decreases SD infringements by 5.7%. An additional level of agreement on a 7-point Likert scale indicating having control over others’ distancing decreases SD infringements by 11.5%. An additional level of agreement on a 7-point Likert indicating feeling normative pressure from neighbours to socially distance increases SD infringements by 5.3%. An additional level of agreement on a 7-point Likert indicating having support from friends increases SD infringements by 13.2%.

### Factors associated with intentional SD infringements

#### Univariate analysis

The differences in means of intentional infringements between categories of each categorical explanatory variable are reported in Table 4. There was a statistically significant difference in intentional infringements between employment status groups as determined by a one-way ANOVA (F(10,670) = 1.86, p= .048). A LSD post hoc test revealed that intentional infringements were statistically significantly higher for self-employed or freelance in normal place of work participants (5.81 ± 5.28) compared to long-term sick or disabled participants (1.32 ± 2.76, p = .002), self-employed or freelance from home (2.03 ± 4, p = .003), working as an employee from home (2.31 ± 4.58, p = .003), unemployed (2.11 ± 3.76, p = .007), working as an employee in normal place of work (2.37 ± 4.07, p = .007), other (2.48 ± 4.15, p = .019), retired (3.09 ± 6.36, p = .037) and looking after home or family (2.97 ± 4.15, p = .047). Intentional infringements were statistically significantly higher for furloughed employees (3.69 ± 5.07) compared to long-term sick or disabled participants (1.32 ± 2.76, p = .023), working as an employee from home (2.31 ± 4.58, p = .031) and self-employed or freelance from home (2.03 ± 4, p = .04). An independent-samples t-test found that participants who were not vulnerable committed statistically significantly more intentional infringements (2.74 ± 4.68) compared to participants who were vulnerable (1.75 ± 4.16), a mean difference of 1 (95% CI, .1 to 1.9), t(149.73) = 2.19, p = .03. There was a statistically significant difference in intentional infringements depending on the lockdown phase relevant to participants’ period of responses as determined by a one-way ANOVA (F(2,678) = 4.12, p= .017). A LSD post hoc test revealed that intentional infringements were statistically significantly higher during the first relaxation phase (3.4 ± 5.56) compared to the full lockdown phase (2.08 ± 4.12, p = .017).

The associations between numerical explanatory variables and intentional infringements are reported in Table 5. There was a moderate, negative correlation between intention to socially distance and intentional infringements (*r_s_*(681) = .409, *p* = .000). There was a weak, negative correlation between knowledge (*r* (681) = -.08, *p* = .036), social responsibility (*r_s_*(681) = -.147, *p* = .000), control over leaving the house, (*r_s_*(681) = -.1, *p* = .009), control over others’ distancing (*r_s_*(681) = -.09, *p* = .019), control over responsibilities (*r_s_*(681) = -.13, *p* = .001), normative pressure from family (*r_s_*(681) = -.189, *p* = .000) and normative pressure from friends (*r_s_*(681) = -.21, *p* = .000) and intentional infringements. There was a weak, positive correlation between self-interest and intentional infringements (*r_s_*(681) = .143, *p* = .000).

#### Multivariate analysis

The negative binomial regression model was statistically significant, χ^2^(57) = 308.916, *p* = .000. A Pearson Chi-Square goodness of fit test returned a value greater than 0.5 (χ^2^ (623) = 1.837), indicating that the model fits the data well. When holding other factors constant, an additional year of age increases intentional SD infringements by 1.7%. Intentional SD infringements increase by 33.2% if an individual is a Christian and decrease by 66.6% if an individual follows a non-specified religion, than if an individual has no religion. Intentional SD infringements increase by 205.1% if an individual is self-employed or freelance in their normal place of work, than if an individual is long-term sick or disabled. Intentional SD infringements increase by 50.8% if an individual is not vulnerable, than if an individual is vulnerable. An additional level of agreement on a 7-point Likert indicating trust in the Government decreases intentional SD infringements by 7.4%. Intentional SD infringements increase by 37.4% if an individual reports on their SD infringements during the relaxation of rules after the first national lockdown, than if an individual reports on their SD infringements during the first national lockdown. An additional correct answer to knowledge about COVID-19 and social distancing decreases intentional SD infringements by 11.9%. An additional level of agreement on a 7-point Likert indicating a sense of social responsibility decreases intentional SD infringements by 14%. An additional level of agreement on a 7-point Likert scale indicating intention to socially distance decreases intentional SD infringements by 31%. An additional level of agreement on a 7-point Likert scale indicating having control over others’ distancing decreases intentional SD infringements by 17.2%. An additional level of agreement on a 7-point Likert indicating feeling normative pressure from friends to socially distance decreases intentional SD infringements by 8.8%. An additional level of agreement on a 7-point Likert indicating feeling normative pressure from neighbours to socially distance increases SD infringements by 10.8%. An additional level of agreement on a 7-point Likert indicating having support from friends increases intentional SD infringements by 16.1%.

#### Summary of quantitate results

The multivariate analysis established that vulnerable health, intention to socially distance, control over others’ distancing, normative pressure from neighbours and support from friends were associated with both total SD infringements and intentional SD infringements. Intentional SD infringements were also associated with religion, employment status, trust in government, lockdown phase, COVID-19 and SD knowledge, social responsibility and normative pressure from friends. Total SD infringements was associated with control over leaving the house, which was not associated with intentional SD infringements.

## Qualitative Results

A total of 32 individuals responded to the interview invitation, and after agreeing a time for a call, two were unavailable, leaving 30 participants’ data for analysis. Interviews lasted a mean of 40 minutes (range 23 - 80 minutes). Demographic characteristics are reported in Table 8. Several factors were identified that shaped the behaviours and intentions of non-adherence, which have been categorised using three levels of the Social Ecological Model: individual, interpersonal and community level. Categories and themes and their relationship to unintentional/intentional non-adherence are presented in Table 9. Interview extracts are provided to illustrate themes.

**Table 8.** Characteristics of the interview sample (n=30)

| Characteristics | n | % |
| --- | --- | --- |
| **Demographic** |  |  |
| Gender |  |  |
| Female | 14 | 47 |
| Male | 14 | 47 |
| Other | 2 | 7 |
| Age |  |  |
| 18-29 | 7 | 23 |
| 30-59 | 14 | 47 |
| 60+ | 9 | 30 |
| Ethnicity |  |  |
| White EG* | 19 | 63 |
| Asian EG | 5 | 17 |
| Black EG | 4 | 13 |
| Mixed EG | 1 | 3 |
| Other EG | 1 | 3 |
| Borough |  |  |
| Enfield | 3 | 10 |
| Islington | 8 | 27 |
| Camden | 4 | 13 |
| Haringey | 9 | 30 |
| Hackney | 4 | 13 |
| Barnet | 2 | 7 |
| Employment status |  |  |
| Working as an employee in normal place of work PT/FT | 3 | 10 |
| Working from home | 13 | 43 |
| A Furloughed employee | 1 | 3 |
| Unemployed | 3 | 10 |
| Retired | 7 | 23 |
| Long-term sick or disabled | 1 | 3 |
| Student | 2 | 7 |
| **Housing & vulnerability** |  |  |
| Living with person of vulnerable health status | 3 | 10 |
| Not living with person of vulnerable health status | 27 | 90 |
| **Health** |  |  |
| Vulnerable | 5 | 17 |
| Not vulnerable | 25 | 83 |

\* EG: ethnic group.

**Table 2:**
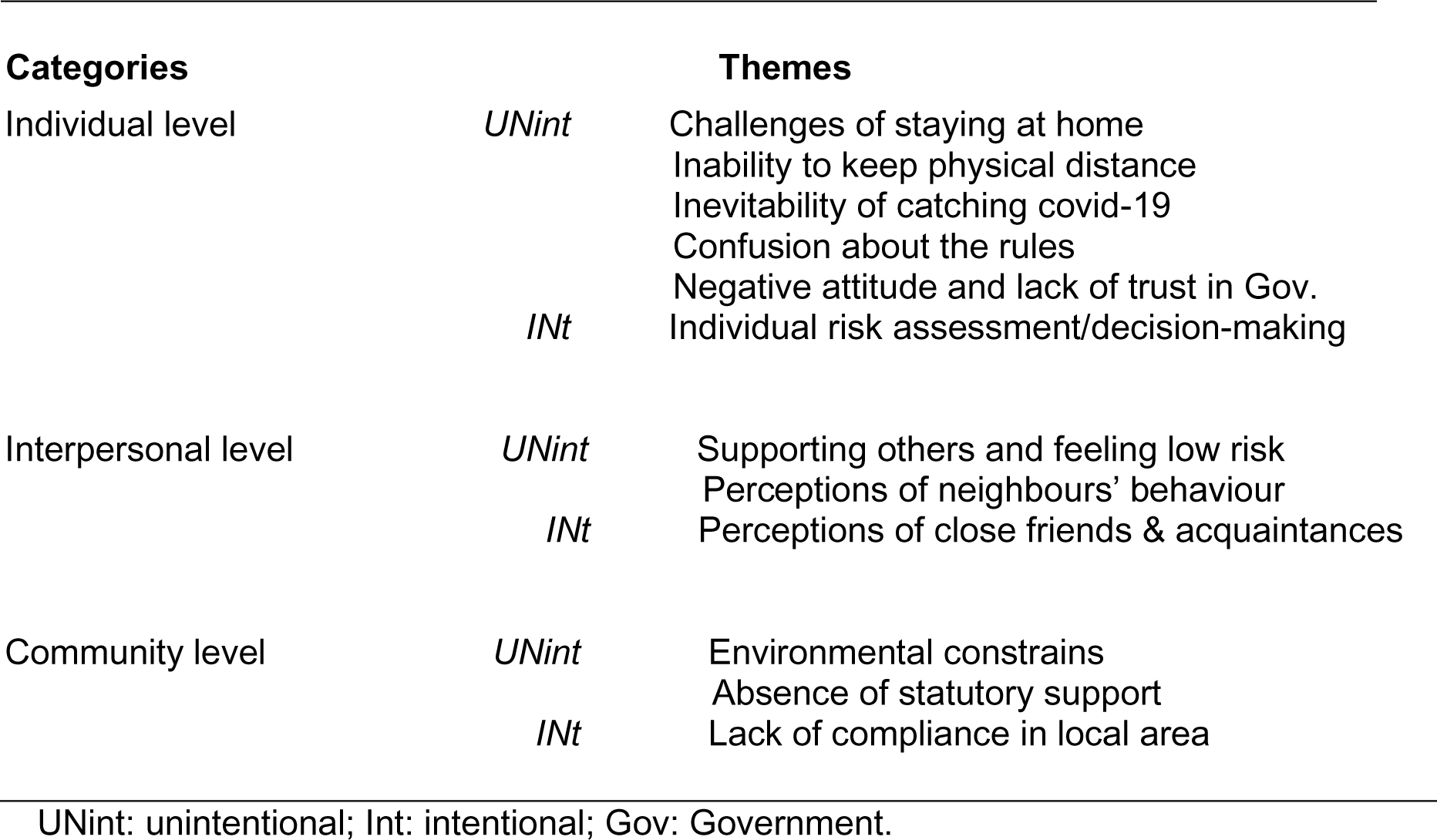
Categorised factors that influence non-adherence to SD rules.

| Categories | Themes |
| --- | --- |

|  |  |  |
| --- | --- | --- |
| Individual level | *UNint* | Challenges of staying at home<br>Inability to keep physical distance<br>Inevitability of catching covid-19<br>Confusion about the rules<br>Negative attitude and lack of trust in Gov. |
|  | *INt* | Individual risk assessment/decision-making |
| Interpersonal level | *UNint* | Supporting others and feeling low risk<br>Perceptions of neighbours' behaviour |
|  | *INt* | Perceptions of close friends & acquaintances |
| Community level | *UNint* | Environmental constraints<br>Absence of statutory support |
|  | *INt* | Lack of compliance in local area |

UNint: unintentional; Int: intentional; Gov: Government.

### Individual level

Individual level factors are related to intrapersonal psychological constructs such as perceived ability and controllability to perform the behaviour as well as attitudes and decision processes towards the rules. Amongst these, the challenges of staying at home, to which many participants found harder than keeping 2 mts. distancing, was associated with perceived difficulties to perform the behaviour (self-efficacy). Participants referred to emotions and personality traits (i.e. feeling anxious, lonely, reclusive, ‘mentally taxing’ or being an extrovert) as well as to house space (living in a small flat or without a garden). This led many men, in particular, and of all ages to leave home for unpermitted reasons, such as exercising more times than it was permitted, or for longer periods, or driving or cycling longer distances than permitted during the full lockdown period. Many argued that they found a ‘safe way’ of doing it, others referred only to the positive impact on their mental health. Someone walking in the day and running in the night, commented: *‘I would have gone crazy if I couldn’t go running’* (M10, WEG, 60s); similarly, another mentioned, ‘*definitely, I had to leave, I couldn’t stay at home’* (M02, BEG, 35+).

On the other hand, inability to keep physical distance was expressed by many as frustrating (lack of controllability) in particular with regard to people’s ‘lack of space awareness’ when shopping or walking on narrow pavements. A few participants who worked outside the home were also concerned about coming into close contact with people. There were also situations of lack of controllability reported by individuals living in shared accommodation for whom leaving the house for exercise and volunteering was a way of regaining self and peer-esteem. One of them commented in relation to his outings:

*‘it kind of got me out of their way [friends] and is quite weird when you’re the only one self-employed and not working. After a month, actually, one of them got furloughed as well so that made it a lot easier, but the other two worked [from home] all the way through lockdown’* (M12, WEG, 25+).

Another individual living with his family and a shielding wife stated they could not follow the SD rules provided for shielding people given the size of their flat.

The majority of informants from the young and adult groups felt they were likely to catch COVID-19, many felt they had caught it previously, a few considered work exposure and living with a key worker as a ‘strong likelihood’, while others thought of the high transmissibility of the virus and contact with people as being ‘inevitable’:

*‘I think I might have had it. I think I might get it, but I am not worried because I am relatively young’* (F14, WEG, 25+).

*‘Yes, I’m likely to catch it. So much unknow about it so everybody can catch it’* (O02, WEG, 40s).

Young people thought that being ‘young, healthy and fit’ made them unlikely to be severely affected by the disease, although a few knew of people who were young and seriously ill. In the adult group, those having mild underlying health issues were concerned about long-COVID (severity); whilst the older group felt less likely to catch COVID-19 due to ‘taking precautions’, and at the same time, many said that it was a ‘lottery’. Those that were clinically vulnerable were more likely to feel both susceptible and to fair worse if caught it, but two mentioned going out during the relaxation of measures, to restaurants, friends’ gatherings, and holydays abroad. One of them weighed up the threat about the virus with feeling lonely, whilst seeking reassurance from friends who had the disease:

*‘I do worry about catching it but I have felt the need for company in the later period in a way that I wasn’t feeling as acutely earlier, and I’ve also felt that because I’ve been reading a lot and sharing information with friends […] who had it [covid] and got through it’* (F09, AEG, 60s).

Other individual factors included confusion about the rules. Informants expressed that their knowledge about the guidelines changed considerably after the first ease of lockdown measures, with most of them stating that they struggled to understand the rules:

*‘Now it’s really hard to know what’s okay to do. I’d like some nice clear information, and clear instruction that wasn’t contradictory. But it’s impossible to get’* (F01, WEB, 70s).

*‘The messaging became very muddle, and almost a joke (‘staying alert’). Friends are not sure of what you can or cannot do. It was confusing and contradictory’* (M11, WEB, 20s).

Confusion about rules also led to negative expressions towards the new rules considered as ‘contradictory’, ‘ridiculous’ and ‘inconsistent’ in the balance of lifting and restricting, especially in regard to norms concerning social interaction (‘I can go to the pub but I can’t meet my friends’; ‘back to work but no seven friends indoors’). At the same time, distrust in the Government’s handling of the pandemic was very high, as with only one exception, who considered the pandemic a new phenomenon and therefore ‘normal’ for the Government to make mistakes, the rest of the participants strongly commented on the Government’s ‘lack of leadership’, ‘complete incompetence’, ‘lack of honesty’ and on their ‘lack of faith’ and ‘lack of trust’. Some of the criticism was directed at the need to impose lockdown earlier, the questioning of an early relaxation of measures, lack of access to tests for symptomatic people, no mandatory use of face masks, no police enforcement, and no border controls, amongst others. Individual decision making, often interchangeably described as following ‘common sense’, performing ‘own risk assessment’, or ‘doing what is sensible to do’ was expressed by many as a personal strategy to counter the Government’s perceived mismanagement of the pandemic:

*‘now I am doing what I feel is right, it seems more like a judgement call than any sort of rule’* (M03, WEG, 25+).

*‘I have breached the guidance in some ways since lockdown ended. I follow my own perceptions on what is safe or not’* (M13, WEG, 35+).

*‘Doing the right thing and complying are not necessary the same thing. I’ve not complied with their [Government] instruction. Right the way through we implemented it [health protection] earlier, so I will continue doing what I feel is right rather than what they suggest’* (F12, BEG, 35+).

*‘We [brothers’ family] did a risk assessment and we decided before the bubble was implemented that we were going to create our own bubble’* (O02, WEG, 40s).

In many cases, this led to a cautious attitude i.e. doing less than it was permitted (not using transport, restaurants and pubs) or, as one of them put it, ‘living in my own little lockdown’. Related to this was a view of reconsidering adherence in the future (intentions), if ‘lockdown’ measures were reinstated. Young and adult groups were more likely to re-consider adherence, some explicitly mentioned rules they would not follow, e.g. if bubbles were suspended, the rule of six, and not meeting people outdoors; others, anticipated an evaluative approach to the measures, similarly to the decision-making process described above. In comparison, older people were more prone to state they would follow the rules in the future.

### Interpersonal level

Interpersonal level factors are those associated with individuals’ perceptions of and interactions with family, friends, and neighbours (social network), as well as support received and provided to facilitate adherence to rules. Young people and adults often expressed prosocial values in terms of feeling socially responsible for helping those most vulnerable given that they perceived themselves as less severely affected if catching Covid-19. Many joined Mutual Aid Groups, others spontaneously offered help to neighbours, often as a response to a perceived lack of support from the Government:

*‘The main thing to come out from this on a local level was that (and I did it myself) I contacted neighbours of mine who are less able to go out and more high-risk categories and asked if they needed any help. And I think that’s what a lot of people did, but that was from a local level … that should have been from a Government level’* (M04, OEG, 35+).

Many in the younger groups who were volunteering admitted to breaking the rules in terms of going out for non-permitted reasons while having COVID-19 symptoms, and at the same time, feeling responsible for the community and wanting to help. In these cases, the latter appears as more important than the perceived risk of spreading the disease to the most vulnerable. One volunteer, for example, commented regarding his approach to rules during lockdown:

*‘the oldest is 28 the youngest was 25 at the time, but, you know, we kind of see ourselves less at risk because we’re not interacting closely with people, we don’t have family around us, we’re not interacting with people more at risk on a regular basis, I guess we then felt that we could kind of follow the rules more loosely’* (M12, WEG, 25+).

Older people displayed more resources in terms of the type of activities they became involved in (Mutual Aid groups, voluntary organisations, providing emotional support over the phone, donations, and cooking). Most valued the work of neighbours supporting the community. On the other hand, adults and older people were more likely to mention the use of masks as protective behaviour than young respondents, and older people considered the risk of running errands if they were living with someone shielding:

*‘We supported people with food. I thought too much of a risk to offer help and drive’* (M14, WEG, 65+).

Neighbours were generally praised on the basis of their support offered to the community, rather than their behaviour towards the rules. There were, however, a few interviewees visibly upset about neighbours constantly flouting the rules, leaving them pondering about their own compliance and self-sacrifice for the collective effort:

*‘it was generally around that age group – maybe more like 20 something, mid-30s from professional backgrounds, but they just.. no compliance whatsoever. That was the only thing that did create animosity, because I am doing all the things that I am supposed to do, but some people out there that don’t give a dam basically, they are countering all the measures. Then you think of course there will be a level of non compliance, but I was quite surprised that it was that group and to that extent’* (M04, OEG, 35+).

Family and friends were mentioned as the main source of emotional support to help staying at home. Remote communication, via video-chat and mobile calls, was used by most participants in all groups. Yet some participants felt the need to meet, in person, members of their family who did not leave with them during the lockdown:

*‘So, I got in the car, I drove 10 minutes. I met him [son] in a park and we were sitting far away from each other but, you know, at least we were able to talk, rather than on Zoom’* (M01, WEG, 70s).

Some also relied upon family and friends for the delivery of food when shielding or having symptoms. Only a few informants received support from neighbours at least once for the delivery of food or prescriptions. Friends were often perceived as being adherent to SD rules, but during relaxation, many referred to creating ‘friend’s rules’ to meet up safely, regardless of the Government guidance.

*‘so we always made sure we met somewhere where everyone could avoid public transport and people, so in that sense, I don’t know if we were compliant at that point, but I felt like we were looking after each other and taking good measures’* (O01, AEG, 30s).

Many interviewees also remarked on ‘close friends’ being compliant as opposed to ‘acquaintances’ or ‘distant friends’, whom they knew were not following the rules and meeting up with others during lockdown.

*‘very intelligent people [other friends] and the fact that they wouldn’t follow the rules on the one hand makes me angry, but then also like makes me go, well, there’s probably a lot of people like them that weren’t following the rules properly, so yeah, it certainly makes it, you know? how you follow the rules in future if there was another lockdown’* (M12, WEG, 25+).

### Community level

Community level captures interactions with local area environments and services as well as perceptions about the local area. There were a range of issues raised in relation to the environment that had a direct relationship with respondents’ ability to comply with SD rules (unintentional non-adherence). The impossibility to access online delivery for foods was a constant problem stated by informants, leaving many of them to find ways of adapting and responding to their new situation, from visiting shops at different hours to changing the way they shopped. Yet despite their strategies, many referred to the lack of enforcement of SD rules by shop owners (groceries, corner shops) for which people turned to for basic items, as well as by supermarkets, where control for protective measures (physical distancing and use of masks) was considered rather weak.

*‘Where I live, there was no social distancing in the small shops […] Some people carried life as normal’* (M02, BEG, 35+).

*‘Supermarket, there wasn’t too much enforcement beyond a few signs’* (M11, WEG, 20s).

Local parks were mentioned as problematic because of being too busy, with groups of people not being dispersed or controlled by the police, and for lack of maintenance; ‘too much litter’. This led people to find alternative green spaces, sometimes having to travel longer distances, or to avoid visiting parks completely:

*‘I didn’t go to the park anymore. Height of lockdown and it was a nice day, it was packed – people playing football and people in the ground. So busy. So, I went to another place/park and was busy as well. Nowhere to distance from people … didn’t see police around at all’* (O02, WEG, 40s).

In addition, restaurants, after they were allowed to open on 4th July, were perceived for some as unsafe, leaving a few regretting their visit. Some indicated this was due to the lack of use of facemasks, others due to the lack of distancing between tables, and the no recording of customers’ contact details, as it was then required. As one commented: *‘I do expect a restaurant to keep my details’* (M09, WEG, 65+).

Regarding support from statutory services, this was very limited for those who needed it, leaving them unable to adhere with the norms as per guidelines. For example, a single mother with underlying health conditions complained about the late distribution and ‘in-store only’ use of food vouchers (free school meals). Likewise, delay in responses was mentioned by those living with a shielding person, where the delivery of food took around a month to get sorted, leaving them feeling at risk when going shopping. A woman with mental health problems and living alone was discharged from hospital at the beginning of mitigation measures and stated it took six weeks for the hospital to get in touch and offer some support. She recalled being scared and non-compliant during that period:

*‘I had it tough because I have mental health issues so it [pandemic] made it worst […] I was discharged from hospital earlier than I should have because of the situation, and I didn’t have the support. I was drop literally into lockdown basically [..] Yes, at the begging I was like no one is gonna force me, no one is gonna police it. I thought, how can you make me stay at home? I’ve got no disease, I know I am disease free. It did take three weeks to sink in’* (F02, AEG, 35+).

Local area perceptions across all age groups indicated a concern about people not complying with SD rules, in particular after the relaxation period. Middle age and older groups considered this an issue with young people gathering in parks or not physically distancing when in shops or in the street. Many informants felt a lack of policing and enforcement of rules as problematic, while others were concerned about a perceived sense of ‘normality’ in the behaviours of local residents:

*‘people don’t wear masks, lots of people are not social distancing. People think is over’* (F01, WEG, 70s).

*‘There is a sense of normality; even in the media referring to ‘during the pandemic’ as if it’s over! That’s interesting language, I think, because I don’t think is over at all.’* (M10, WEG, 60s).

## Discussion

This study identified individual, interpersonal and community level factors associated with intentional and unintentional non-adherence to SD rules during the first ‘lockdown’ and relaxation in a cross-sectional North London sample. To our knowledge, this is the first mixed-methods study on SD measures combining a survey with in-depth interviews. The explanatory sequential design allowed for an integration of qualitative data to offer a deeper understanding of quantitative findings whilst shedding light on new emerging themes about people’s non-adherence behaviours. It is important to consider that the relaxation period introduced significant changes to SD rules and that interviews took place between two and three and a half months after completion of the survey.

The additional detail gained from the qualitative data demonstrates aspects that are unique to a population group that could be broadly defined as being able to stay at home (able to work from home or retired), with lower caring responsibilities outside the house, largely not vulnerable to COVID-19, and who were exceptionally relying on statutory services.

Participants’ accounts revealed different challenging situations experienced both during full ‘lockdown’ and the relaxation of rules at the intersections of individual, interpersonal and community levels. The most frequent act of non-adherence, as reported in the survey (on average 1.92 times) and interviews, was going out for not permitted reasons (time and frequency of exercising and not staying local), often in relation to psychological factors (self-efficacy) that affected their confidence to staying at home i.e. feeling confined. The second most reported act of non-adherence in all groups was at the interpersonal level; meeting family or friends they did not live with (at least once during lockdown and more than once during relaxation), as initially identified in our survey findings (average 0.67). Survey data reported that an additional level of support from friends increased the odds of intentionally not adhering to SD rules by 16%, which is further explained in the qualitative data where participants felt a strong emotional support from friends and family, and justified breaking rules due to ‘missing social interaction’ with loved ones, more likely reported after the relaxation. Indeed, some participants stated that groups of friends agreed to meet up under certain rules (decision-making processes) which meant a contravention of guidelines. The perceived adverse emotional impact of SD rules and non-adherent behaviours has been reported in previous reviews [28] and studies [8]. It is worth noting that a study from Quebec [29] found emotional factors as a non-significant predictor of adherence to SD norms, but it did find independent predictors of adherence in civic duty (protecting others), protecting the vulnerable, and social norms (others respecting the rules). Whilst a belief in civic duty relies upon consistent messages and trust in governments, lack of trust may affect the relative influence of other psychological factors, leading to a breakdown in known behavioural associations such as a perceived threat and response efficacy (believing measures are effective) in the adoption of protective behaviours [30]. In our qualitative sample, participants largely believed SD measures were effective on their own, but the handling from the Government, especially with regard to the timing of its implementation (i.e. too late for full lockdown, too early for relaxation) alongside other factors discussed below, led behaviours to be informed by individual risk assessments/decision-making, rather than ‘protecting others’.

Our multivariate analysis identified that an additional level of agreement on a 7-point Likert scale indicating trust in the Government decreases intentional SD infringements by 7.4%. The qualitative data enabled us to identify two possible explanations for this. First, attitude (emerging theme – not queried in the survey) revealed a negative perception towards the management of SD rules, in particular, after relaxation, which was informed by a lack of trust in Government, confusion about the content of the rules (knowledge), and a perception of conflicting messages, i.e. contradiction between what was permissible to do and not. Second, this negative attitude led, in turn, to a decision-making process (emerging theme) whereby individuals reported making their own evaluation about what was ‘reasonable’ or ‘safe’ to do in a particular context. The interaction of these variables allowed us to identify this as a key explanatory factor, where lack of trust in the Government and confusion about rules informed a negative attitude and induced an individual decision-making process where, in many instances, intentional non-adherence took place. These behaviours remind us of the importance of governments in maintaining confidence and public trust in their management of the pandemic, yet evidence indicates that UK politicians have foundered in this key area with people’s trust declining from 66% by mid-April 2020 to 39% in early June and to 30% by mid-September [31].

There was a considerable level of support provided by interviewees to their communities, through engagement with Mutual Aids groups or running errands for neighbours, especially during the lockdown period. Interestingly, for the young group, this reflected an increased sense of community and prosocial values (social responsibility) although, at the same time, we found that this was not always associated with ‘protecting the most vulnerable’, as some volunteers reported going out for non-essential reasons and being symptomatic at different points. This is similar to other UK surveys where low adherence was associated with people engaged in community work while having COVID-19 symptoms [8] or not [32].

At the community level, we were able to identify the main reasons that affected one of the variables in relation to unintentional non-adherence in our survey (control over others distancing, reported on average 8.18 times). Most of the informants reflected on their inability to keep 2 mts. distancing from people when shopping in local convenience stores and supermarkets. Parks were also identified as overcrowded areas, including lack of maintenance and policing to disperse groups. Environmental constrains also included restaurants and pubs, which were questioned for their lack of observance of the rules (wearing masks, contact tracing, distancing in indoors) leaving many feeling unsafe and unintentionally breaking the 2 mts. rules. In addition, there was clearly a reported negative perception of people’s lack of compliance in the ‘local area’, also found in a study from Islington and Camden [33]. Our qualitative data suggested that participants more often perceived lack of adherence in the ‘local area’ in comparison to ‘neighbours’, thus resonating with studies arguing for the need to consider the spatial proximity variance in behaviours affected by social norms [34]. This may explain a counterintuitive finding observed in our survey indicating that an increased normative pressure from neighbours was associated with intentional non-adherence to SD rules. Furthermore, local area lack of compliance (not queried in the survey) may also influence intentional non-adherence behaviours, as other studies have reported [29, 35], and similarly in other social situations when adherence to a required behaviour is conditional on others doing the same.

## Implications

Building trust in public health measures might be challenging for the implementation of future SD measures or other protective measures, such as vaccinations, whose effectiveness depends on people’s behaviours. Reports have shown [36–39] that the UK Government’s centralised management of the pandemic, from decision-making and communication to the roll out of test and trace, has undermined engagement with local health authorities and communities. Partnerships with local communities, early development of strategies for community participation, and then frequent priority needs assessments to strengthen community engagement and support could provide communities with the tools to comply with the guidelines, whilst increasing acceptability. [Factors addressed: lack of trust in Government; individual decision-making; absence of statutory support].

Parallel to partnership working, public health messaging should emphasise prosocial behaviours and responsibility for the community, especially protecting the vulnerable, tailoring messages to young adults who may underestimate the risk of spreading the disease. [Factors addressed: Supporting others and feeling low risk].

It is also recommended that Environmental Health officers should work with local small businesses (corner shops), pubs and restaurants to advise on SD practices and practical protective behaviours, ensuring they are COVID-secure. Whilst it is important to do so by engaging with businesses’ concerns, it is also relevant that environmental health officers have enforcement powers to close business when failing to comply. [Factors addressed: Environmental constrains and lack of compliance in local area].

## Conclusions

Our findings have identified ways in which non-adherence to SD rules can be intentional and unintentional and can be underpinned by factors occurring at different levels; individual, interpersonal, and community. In particular, our findings highlighted the importance of factors beyond people’s control and others that involved conscious decision-making, both resulting in a reduced likelihood to adopt recommended behaviours. Whilst all these factors are modifiable, our findings indicate that non-adherence can be improved by partnerships with local communities to build trust in SD measures; tailored messaging to young adults emphasising the need of protecting others whilst clarifying the risk of spreading the virus; and by ensuring COVID-secured environments through the work of environmental health officers.

## Strengths and limitations

The mixed-methods approach allowed a nuanced understanding by integrating a predictive quantitative model with inferences from participants’ past experiences of behavioural non-adherence. However, there are known limitations to this type of research design. In particular, the challenge of attaining true integration over two large data sets. To overcome this barrier, our qualitative data was derived from the findings of the quantitative survey, so it was possible to integrate the two data sets and findings together by organising it into the different levels of the Social Ecological Model that guided our theoretical approach for both sets of data collection.

Our study was restricted to those living in North London boroughs and behavioural adherence of people living in other parts of London and England might have been different. Further, although our qualitative sample offered a more balanced representation of population groups, in the survey, men and BAME groups were underrepresented, thus caution is needed in generalising quantitative findings to the North London population of interest.

Self-report surveys and interviews are subjected to biases including recall bias (forgetting about breaking the rules) and social desirability bias (not admitting to breaking the rules). Potential interviewer bias was mitigated using a reflexive interviewing approach, whereby honesty in the responses provided was encouraged through a non-judgemental attitude, by ensuring participants understood the questions being asked and the interviewer double checking responses at the end, when necessary.

## Data Availability

The datasets used and/or analysed during the current study are available from the corresponding author on reasonable request.

## Acknowledgments

The authors thank all the participants who completed the survey and the interviews. We acknowledge the following people for their contribution to the development of data collection instruments: Chris Chandler, Olive McKeown, Nirmala Lee, Emma Whitby, Rosa Lau, and Alexandra Watson.

## Notes

### Competing Interest Statement

The authors have declared no competing interest.

### Funding Statement

This study was funded by the Transformation Fund (Higher Education Innovation Fund), London Metropolitan University.

## References

1. Public Health England. Guidance on social distancing for everyone in the UK. 2020 March 30. [cited 2020 Sep 15]. In GOV.UK [Internet]. Available from: https://www.gov.uk/government/publications/covid-19-guidance-on-social-distancing-and-for-vulnerable-people/guidance-on-social-distancing-for-everyone-in-the-uk-and-protecting-older-people-and-vulnerable-adults

2. Brauner JM, Mindermann S, Sharma M, Johnston D, Salvatier J, Gavenčiak T, et al. Inferring the effectiveness of government interventions against COVID-19. Science. 2021 Feb 19;371,6531, eabd9338eabd9338. doi:10.1126/science.abd9338.

3. Public Health England. Coronavirus (Covid-19) in the UK. Cases in United Kingdom. 2020 [Cited 7 February 2021]. In GOV.UK-PHE [Internet] Available from: https://coronavirus.data.gov.uk/details/cases

4. Ipsos MORI. Coronavirus polling October 2020 [Cited 5 March 2021] Available from: https://www.ipsos.com/sites/default/files/ct/news/documents/2020-10/following_coronavirus_rules.pdf

5. Fancourt D, Bu F, Mak HW, Steptoe A. Covid-19 Social Study Results Release 8. 2020 May 13 [Cited 1 March 2021] Available from: https://b6bdcb03-332c-4ff9-8b9d-28f9c957493a.filesusr.com/ugd/3d9db5_1806a0f44fc145bfae64ab7567413bff.pdf

6. Duffy B, Allington D. The accepting, the suffering and the resisting: the different reactions to life under lockdown. The Policy Institute, Kings College London. 2020 April 27 [Cited 2021 March 15] Available from: https://www.kcl.ac.uk/policy-institute/assets/Coronavirus-in-the-UK-cluster-analysis.pdf

7. Atchison C, Bowman LR, Vrinten C, et al. Early perceptions and behavioural responses during the COVID-19 pandemic: a cross-sectional survey of UK adults. BMJ Open 2021 Jan 4;11:e043577. doi:10.1136/bmjopen-2020-043577.

8. Smith LE, Amlȏt R, Lambert H, Oliver I, Robin C, Yardley L, Rubin GJ. Factors associated with adherence to self-isolation and lockdown measures in the UK: a cross-sectional survey. Public Health. 2020 Oct; 187: 41–52. doi: 10.1016/j.puhe.2020.07.024.

9. Smith LE, Potts HWW, Amlôt R, Fear NT, Michie S, Rubin GJ. Adherence to the test, trace and isolate system: results from a time series of 21 nationally representative surveys in the UK (the COVID-19 Rapid Survey of Adherence to Interventions and Responses [CORSAIR] study). [Preprint] 2020 [cited 2021 April 1]. Available from: https://www.medrxiv.org/content/10.1101/2020.09.15.20191957v1.

10. McLeroy KR, Bibeau D, Steckler A, Glanz K. An Ecological Perspective on Health Promotion Programs. Health Educ Q. 1988;15(4):351–377.

11. Hills S, Eraso Y. Factors associated with non-adherence to social distancing rules during the COVID-19 pandemic: a logistic regression analysis. BMC Public Health. 2021 Feb 13;21(1):352. doi: 10.1186/s12889-021-10379-7.

12. Creswell JW, and Plano Clark VL. Designing and conducting mixed methods research. 3rd ed. Thousand Oaks, CA: SAGE Publications; 2017.

13. Office for National Statistics. All data related to Population estimates for the UK, England and Wales, Scotland and Northern Ireland: mid-2018. 2019 June 26 [cited 2020 Sep 16] In Office for National Statistics [Internet] Available from: https://www.ons.gov.uk/peoplepopulationandcommunity/populationandmigration/populationestimates/bulletins/annualmidyearpopulationestimates/mid2018/relateddata

14. Cochran WG. Sampling Techniques. 2nd ed. New York: Wiley; 1963.

15. Ajzen, I. Perceived behavioral control, self-efficacy, locus of control, and the theory of planned behavior. J Appl Soc Psychol. 2002;32: 665–683.

16. Office for National Statistics. Testing the census. 2019 [cited 1 April 2021]. In: Office for National Statistics [Internet]. Available from: https://www.ons.gov.uk/census/censustransformationprogramme/testingthcensus.

17. Ministry of Housing Communities and Local Government. English indices of deprivation 2019: Postcode Lookup. [cited 26 Mar 2021]. In: Ministry of Housing Communities and Local Government [Internet] Available from: http://imd-by-postcode.opendatacommunities.org/imd/2019

18. Gregory TA, Wilson C, Duncan A, Turnbull D, Cole SR, Young G. Demographic, social cognitive and social ecological predictors of intention and participation in screening for colorectal cancer. BMC Public Health. 2011 Jan 14;11:38. doi: 10.1186/1471-2458-11-38.

19. Oosterhoff B, Palmer C. Psychological Correlates of News Monitoring, Social Distancing, Disinfecting, and Hoarding Behaviors among US Adolescents during the COVID-19 Pandemic. JAMA Pediatr. 2020;174(12):1184–1190. doi:10.1001/jamapediatrics.2020.1876.

20. Zimet GD, Dahlem NW, Zimet SG, Farley GK. The Multidimensional Scale of Perceived Social Support. J Pers Assess. 1988 Jun 10;52(1):30–41.

21. Ritchie J, Spencer L. Qualitative data analysis for applied policy research. In: Huberman M, Miles M, editors. The Qualitative Researcher’s Companion. London: Sage; 2002. pp 305–329.

22. Fetters MD, Curry LA, Creswell JW. Achieving Integration in Mixed Methods Designs—Principles and Practices. Health Serv Res. 2013; 48(6 Pt 2): 2134–2156. doi: 10.1111/1475-6773.12117

23. Curtin R, Presser S, Singer E. The Effects of Response Rate Changes on the Index of Consumer Sentiment. Public Opin Q. 2000 Feb 1;64(4):413–28. doi:10.1086/318638.

24. Smith G. Does gender influence online survey participation?: A record linkage analysis of university faculty online survey response behavior. Scholarworks.sjsu.edu. 2008. Available from: https://scholarworks.sjsu.edu/elementary_ed_pub [cited 2021 March 3].

25. Office for National Statistics. Regional ethnic diversity - GOV.UK Ethnicity facts and figures. In Office for National Statistics [Internet]. 2020 [Cited 3 February 2021]. Available from: https://www.ethnicity-facts-figures.service.gov.uk/uk-population-by-ethnicity/national-and-regional-populations/regional-ethnic-diversity/latest

26. Ahlmark N, Algren MH, Holmberg T, Norredam ML, Nielsen SS, Blom AB, et al. Survey nonresponse among ethnic minorities in a national health survey-A mixed-method study of participation, barriers, and potentials. Ethn Health. 2015;20:6: 611–632. doi: 10.1080/13557858.2014.979768.

27. Treweek S, Forouhi NG, Narayan KMV, Khunti K. COVID-19 and ethnicity: who will research results apply to? Lancet. 2020 27 June-3 July; 395(10242): 1955–1957. doi: 10.1016/S0140-6736(20)31380-5

28. Teasdale E, Santer M, Geraghty AWA, Little P, Yardley L. Public perceptions of non-pharmaceutical interventions for reducing transmission of respiratory infection: systematic review and synthesis of qualitative studies. BMC Public Health. 2014;14:589 Available from: http://www.biomedcentral.com/1471-2458/14/589

29. Gouin J, MacNeil S, Switzer A, Carrese-Chacra E, Durif F, Knäuper B, et al. (2021). Socio-demographic, social, cognitive, and emotional correlates of adherence to physical distancing during the COVID-19 pandemic. Can J Public Health. 2021 Jan 19. https://doi.org/10.17269/s41997-020-00457-5.

30. Rippetoe PA, Rogers RW. Effects of components of protection-motivation theory on adaptive and maladaptive coping with a health threat. J Pers Soc Psychol. 1987 Mar;52(3):596–604. doi: 10.1037//0022-3514.52.3.596.

31. YouGov. YouGov COVID-19 tracker: government handling. 2021 Mar 17 [Cited 5 March 2021] Available from: https://yougov.co.uk/topics/international/articles-reports/2020/03/17/perception-government-handling-covid-19

32. Wright L., Steptoe A, and Fancourt D. (2020) What predicts adherence to COVID-19 government guidelines? Longitudinal analyses of 51,000 UK adults. Preprint medRxiv 2020.10.19.20215376; doi: 10.1101/2020.10.19.20215376.

33. Camden and Islington Public Health Knowledge, Intelligence and Performance team. Covid 19 Resident Engagement Full Report. Findings from resident survey, focus groups and insights gathered by both councils & VCS partner organisations. October 2020. [cited 27 March 2021] In: Islington Council [Internet] Available from: https://www.islington.gov.uk/-/media/sharepoint-lists/public-records/publichealth/information/adviceandinformation/20202021/20201029covid19residentengagementfullreportfinal281020.pdf

34. Passafaro P, Livi S and Kosic A. Local Norms and the Theory of Planned Behavior: Understanding the Effects of Spatial Proximity on Recycling Intentions and Self-Reported Behavior. Front. Psychol. 2019;10:744. doi: 10.3389/fpsyg.2019.00744.

35. Coroiu A, Moran C, Campbell T, Geller AC. Barriers and facilitators of adherence to social distancing recommendations during COVID-19 among a large international sample of adults. PLoS One. 2020 Oct 7; 15(10): e0239795. doi: 10.1371/journal.pone.0239795.

36. C19 National Foresight Group, Nottingham Trent University. Covid-19 Pandemic: Third Interim Operational Review (September 2020). [cited 1 April 2021]. In: C19 National Foresight Group [Internet] Available from: https://www.ntu.ac.uk/__data/assets/pdf_file/0027/1177902/NTU-C19-NFG-Report-0920-Communications-and-the-Covid-19-Pandemic-Rapid-Report.pdf

37. Dyer C. Covid-19: UK government response was overcentralised and poorly communicated, say peers. BMJ 2020; 371 :m4445. doi:10.1136/bmj.m4445.

38. UK Parliament. Biosecurity and national security Contents. 4 Resilience on the ‘frontlines’. 2020 Dec 18 [cited 1 April 2021]. In UK Parliament [Internet] Available from: https://publications.parliament.uk/pa/jt5801/jtselect/jtnatsec/611/61107.htm

39. Independent SAGE. COVID-19: what are the options for the UK? Recommendations for government based on an open and transparent examination of the scientific evidence. May 12 2020 [cited 2 April 2021]. In Independent SAGE [Internet] Available from: https://www.independentsage.org/wp-content/uploads/2020/05/The-Independent-SAGE-Report.pdf

